# Predicting neuropsychological testing outcomes and research clinic diagnosis of MCI and dementia in Parkinson’s disease using the MoCA

**DOI:** 10.1101/2025.06.30.25330565

**Authors:** Tanja Zerenner, Sanjay G Manohar, Jamil Razzaque, Falah Al Hajraf, Karolien Groenewald, Ludo van Hillegondsberg, Jessica Welch, Tamir Eisenstein, Daniel Weintraub, Johannes Klein, Yoav Ben-Shlomo, Michele T Hu, Michael Lawton

## Abstract

**Background and Objectives:** The Montreal Cognitive Assessment (MoCA) is frequently used in cohort studies in Parkinson’s disease (PD) as a simple and quick tool for assessing global cognitive abilities of patients. However, cut-off values for distinguishing between normal cognition, mild cognitive impairment (MCI), and dementia differ across the literature. We comprehensively evaluate the accuracy of the MoCA for patient stratification and whether it can improved by including additional routinely collected information.

**Methods:** We use longitudinal data from PD and healthy control (HC) participants of the PPMI cohort which – in addition to the MoCA – conducts detailed neuropsychological testing and records diagnoses of MCI and dementia made at the research clinics. Multilevel logistic regression was used to predict (1) impairment in detailed neuropsychological testing and (2) clinician diagnoses from the MoCA in conjunction with other routinely collected information on basic demographics or functional impairment as recorded in MDS-UPDRS 1.1. Model performance was assessed using the area under the ROC curve (AUC). Optimal cut-offs for patient stratification were derived according to Youden’s J, common screening and diagnosis criteria, and an equal proportions criterion, that is, the cutoff at which the proportion of observations with the outcome equals the proportion of observations below cutoff.

**Results:** We analysed data from 1,094 PD patients and of 267 HC. Education-adjusted MoCA scores predicted impairment in 2 or more domains with an AUC of 0.86 (95% CI 0.84, 0.88). Youden’s J was maximized at cutoff ≤ 24 with sensitivity 74.7 (70.5, 79.3) and specificity 83.1 (82.0, 84.2); cutoff ≤ 21 equated proportions. The MDS-UPDRS 1.1 was a better predictor of research clinic diagnosis of PD-MCI or PDD than the MoCA. A combination of MDS-UPDRS 1.1 and education adjusted MoCA discriminated diagnosis of any impairment from no impairment with an AUC of 0.87 and dementia from no dementia with an AUC of 0.96.

**Discussion:** Optimal MoCA cutoffs for PD-MCI or PDD depend on their purpose. For post-hoc stratification for research purposes, we recommend considering a cutoff that equates proportions. Identifying suitable cutoffs from the literature - for research or in clinic use - needs to take into account the respective PD population.

## 1 Introduction

About 25% of PD patients with normal cognition at the time of PD diagnosis convert to mild cognitive impairment (MCI) within 3 years [1]. The annual risk of dementia in a PD prevalent population was estimated as 4.5% in a recent meta-analysis [2]. An accurate assessment of the cognitive state of PD patients is crucial not only in clinic, but also for research purposes, for example for identifying risk factors for cognitive decline and future dementia [2, 3], predictive modeling [4, 5], or biomarker research [6].

Ideally a patient’s cognitive state is assessed with an extensive test battery comprised of neuropsychological tests for all cognitive domains. In Parkinson’s neuropsychology a test battery comprising two tests for each of the 5 major cognitive domains is referred to as a ‘Level-II’ assessment [7]. Such detailed neuropsychological testing is however time-consuming and can only be administered by a trained psychophysiologist. Therefore, it is in practice often not feasible and either reduced test batteries or so-called ‘bedside’ tests, which are quick and can be administered without specific training, are utilized instead.

One of the currently most widely used test is the Montreal Cognitive Assessment (MoCA), a 10 min test comprising 30 questions assessing executive and visuospatial function, naming, attention, language, abstraction, recall, and orientation [8]. It is widely acknowledged that the MoCA has not been specifically designed for the profile of cognitive impairment specific to PD and hence cutoffs optimal in the general population are not necessarily optimal in a PD population. Numerous studies have thus addressed the derivation of PD specific MoCA cutpoints for PD-MCI and/or PDD [9–21]. Optimal cutoffs reported vary from ≤ 17 to ≤ 26 for PD-MCI and ≤ 9 to ≤ 25 for PDD depending on purpose (e.g., screening vs diagnosis), optimality criteria (e.g., Youden’s J, balance of sensitivity and specificity) or study population.

We evaluate the ability of the MoCA screening to predict the results of detailed neuropsychological testing as well as research clinic diagnoses of PD-MCI or PDD using longitudinal data from over 1,000 study participants diagnosed with PD. Furthermore, we investigate if predictions can be improved by incorporating additional routinely collected information on basic demographics, functional impairment (MDS-UPDRS 1.1) or by optimizing the weighting of MoCA subscores. Finally, we consider four different criteria for selecting optimal cutoffs for research and in-clinic use and report respective cutoffs for the best model/covariate combination.

## 2 Methods

### 2.1 Data

We utilzed data from the the Parkinson’s Progression Markers Initiative (PPMI) cohort study. Details on recruitment, inclusion/exclusion criteria and follow-up are described elsewhere https://www.ppmi-info.org [22, 23]. Data was downloaded from the PPMI data portal on 12th July 2024 https://www.ppmi-info.org/access-data-specimens/download-data.

Routine variables and assessments collected in the PPMI cohort include basic participant demographics, the MDS-Unified Parkinson’s Disease Rating Scale (MDS-UPDRS) [24] and the MoCA [8]. Part 1 of the MDS-UPDRS assesses non-motor experiences of daily living/question 1 assesses functional impairment due to cognitive deficits as perceived by the patient and/or caregiver on a scale from 0 (no functional impairment) to 4 (severe functional impairment). The MoCA total scores were adjusted for education by adding one point for individuals with ≤ 12 years of education (up to a maximum of 30).

In the PPMI cohort detailed neuropsychological testing of executive function, memory and visuospatial function has been conducted since the study began in 2010. More recently the test battery has been extended, but due to the comparably low number of complete observations for the new, extended battery, we restrict our analysis to initial battery collected since study begin: Memory has been assessed with the Hopkins Verbal Learning Test - Revised (HVLT-R)[25] of which we consider the scores for immediate recall and recognition discrimination. Executive function has been assessed in three distinct tests, animals fluency test [26], Symbol Digit Modalities Test (SDMT) [27, 28] and Letter-Number-Sequencing (LNS) [29]. Visuospatial functioning has been assessed with the 15-item (odd/even) version of the Benton Judgment of Line Orientation (BJL) [30].

We refer to the “cognitive categorization” from the PPMI protocol as “research clinic diagnosis”. A diagnosis of PD with mild cognitive impairment (PD-MCI) is made if there exists (i) a decline from prior usual cognitive abilities, (ii) impairment in at least one cognitive domain, but (iii) no significant impact of on daily functioning. A diagnosis of PD dementia (PDD) is made if there exists (i) a decline from prior usual cognitive abilities, (ii) impairment in more than one cognitive domain, and (iii) significant impact of cognitive impairment on daily function. To determine the categorization, the site investigator may review cognitive test performance, consider the degree of functional impact reported by the participant, a caregiver/informant or observed during the research clinic as well as their overall impression of the participant’s cognitive abilities.

### 2.2 Evaluating neuropsychological testing

Initially we attempted to evaluate the neuropsychological testing data utilizing normative ranges from existing literature, but we found the percentages of patient visits with test scores < 1.5 standard deviations (SD’s) below norm to vary greatly across the different tests when applying respective published norms. For the BJL, for example, the percentage of PD < 1.5 SD below norm [31, 32] was less than the 6.5% expected in a healthy population. Similar was recently reported by [33].

We therefore opted to utilize the longitudinal neuropsychological testing data available for the controls arm of the PPMI cohort to derive our own age-, sex- and education-adjusted normative ranges for LNS, SDMT, animals fluency, BJL, HVLT-immediate recall and HVLT-discrimination using mixed-effects linear and tobit regression. The latter was used if floor or ceiling effects were evident in the statistical distribution of the test scores. Details are provided in the Supplementary Material.

To quantify test performance in each cognitive domain, the age, sex and education adjusted z-scores of the individual tests are summed within each domain:

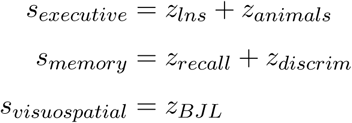

We chose to not include the SDMT in the executive domain score *s_executive_* as it appears to be impacted by motor function more strongly than the other scores.

The domain level scores *s_exeutive_* and *s_memory_* are no longer z-scored, but have a standard deviation < 1 due to the positive correlation of the individual test scores. Instead of z-scoring a second time we opted to use the 6.5%-quantiles of the healthy controls domain scores as a cutoff for impairment in the respective domain (Supplementary Table 4). In a standard normal distribution the 6.5%-quantile corresponds to −1.5 SD’s below expectation/norm. Hence our cut-off resembles one of the criteria routinely applied in the literature to define impairment in neuropsychological testing as −1.5 SD’s below norm [34, 35].

### 2.3 Statistical modeling

We consider a total of four different outcomes: (1) impairment in ≥ 1 domain in detailed neuropsychological testing (2) impairment in ≥ 2 domains, (3) research clinic diagnosis of cognitive impairment (PD-MCI/PDD vs. PD-NC) and (4) research clinic diagnosis of dementia (PDD vs. PD-NC/PD-MCI). We first assess the discriminative ability of the MOCA total scores. We then proceed to study if discriminative ability can be improved by (a) utilizing MoCA subscores instead of totals, (b) incorporating routinely collected information on basic demographics (age, sex and years of education) or (c) incorporating functional impairment as assessed in MDS-UPDRS 1.1. The MoCA subscores we consider are based on the categorization of questions on the MoCA instrument, i.e., visuospatial/executive, naming, attention, language, abstraction, delayed recall, and orientation [8, 36]. The MDS-UPDRS 1.1 responses are included as a factor variable with categories 3 (moderate) and 4 (severe) merged into a single factor due to 4 being rarely recorded (only 10 times in a total 4700 patient visits, 0.2%, Table 1). For the research clinic diagnosis outcomes we further study the discriminative ability of the neuropsychology domain scores, i.e., *s_exeutive_*, *s_memory_* and *s_visuospatial_* in comparison to the MoCA.

**Table 1:** Demographics and cognition in the PD arm of the PPMI cohort stratified by the number of domains impaired according to neuropsychological testing and by research clinic diagnosis. The table summarizes the longitudinal visits from 1094 different patients, of whom 1086 received a diagnosis from the research clinic at least once during their visits. Shown are means and standard deviations or, where applicable, percentages and counts.

|  | impaired domains (neuropsychology) |  |  |  | research clinic diagnosis |  |  |
| --- | --- | --- | --- | --- | --- | --- | --- |
|  | 0 | 1 | 2 | 3 | PD-NC | PD-MCI | PDD |
| N [patient visits] | 3431 (73%) | 881 (18.7%) | 297 (6.3%) | 91 (1.9%) | 3545 (77.8%) | 910 (20.0%) | 104 (2.3%) |
| age | 65.1 (9.4) | 66.5 (10.2) | 68.5 (10.4) | 67.3 (10.9) | 64.5 (9.5) | 70.0 (8.5) | 72.4 (9.2) |
| female | 37.5% (1286) | 40.2% (354) | 35.7% (106) | 38.6% (35) | 40.1% (1420) | 31.2% (284) | 32.7% (34) |
| education years | 15.9 (3.2) | 15.3 (3.5) | 14.8 (4.2) | 14.3 (4.7) | 15.8 (3.4) | 15.5 (3.3) | 14.7 (4.3) |
| PD duration [years] | 5.4 (3.3) | 5.4 (3.3) | 6.2 (3.5) | 6.5 (3.3) | 5.3 (3.2) | 6.6 (3.5) | 7.6 (3.1) |
| MoCA total (adj)** | 27.4 (2.4) | 25.3 (3.2) | 22.4 (4.0) | 18.0 (4.8) | 27.1 (2.6) | 24.6 (3.7) | 19.4 (5.1) |
| MDS-UPDRS 1.1 |  |  |  |  |  |  |  |
| 0 (none) | 65.5% (2244) | 50.3% (442) | 37.4% (111) | 15.4% (14) | 71.5% (2532) | 19.6% (178) | 4.8% (5) |
| 1 (slight) | 27.7% (948) | 34.1% (299) | 34.7% (103) | 39.6% (36) | 25.9% (916) | 47.0% (427) | 10.6% (11) |
| 2 (mild) | 5.6% (192) | 11.5% (101) | 16.9% (50) | 25.3% (23) | 2.4% (85) | 26.1% (237) | 36.5% (38) |
| 3 (moderate) | 1.2% (40) | 3.8% (33) | 10.4% (31) | 14.3% (13) | 0.3% (10) | 7.2% (65) | 40.4% (42) |
| 4 (severe) | 0.0% (0) | 0.3% (3) | 0.7% (2) | 5.5% (5) | 0.0% (0) | 0.2% (2) | 7.7% (8) |
| PD-NC | 85% (2829) | 67.8% (576) | 41.9% (121) | 20.9% (19) | - | - | - |
| PD-MCI | 14.5% (484) | 29.7% (252) | 44.6% (129) | 49.5% (45) | - | - | - |
| PDD | 0.5% (17) | 2.5% (21) | 13.5% (39) | 29.7% (27) | - | - | - |
| impaired domains<br>(neuropsychology) |  |  |  |  |  |  |  |
| 0 | - | - | - | - | 80.0% (2827) | 53.3% (484) | 16.4% (17) |
| 1 | - | - | - | - | 16.3% (576) | 27.6% (251) | 20.2% (21) |
| 2 | - | - | - | - | 3.5% (121) | 14.2% (129) | 37.5% (39) |
| 3 | - | - | - | - | 0.5% (19) | 4.9% (45) | 26.0% (27) |

\*\* The MoCA scores were adjusted for education by adding one point for individuals with $\leq 12$ years of education (up to a maximum of 30).

As we use longitudinal data and therefore non independent observations, we employ mixed-effect logistic regression with a random intercept for each patient. As we aim to derive models for estimating outcome probabilities of ‘new’ patients for whom no random effect was estimated, we need to obtain and evaluate marginal predictions. Due to the non-linear logit link function of the logistic regression, we cannot simply apply the conditional model with the random effect set to zero [37, 38]. Marginal effects *β_M,m_*, however, can be estimated from the respective conditional effects *β_RE,m_* using a cumulative Gaussian approximation to the logistic function [38, 39]. Details are provided in the Supplementary Material. One can then proceed to estimate the probability of an outcome for a ‘new’ patient utilizing the marginal effects *β_M,m_*, like in a standard, i.e., non-multilevel, logistic regression as

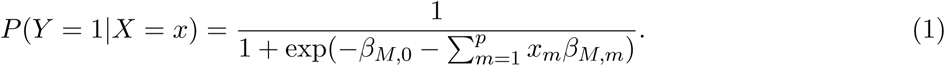

Models were fitted using R’s *lme4* package [40] with the *nlminbwrap* optimizer. ROC curves were computed and plotted using *pROC* [41]. Calibration curves were plotted with the *CalibrationCurves* package. Following recommendations for internal validation of logistic regression prediction models [42], we performed a non-parametric bootstrap. Bootstrap estimation of bias/model optimism allows for optimism correction of the AUC estimates. The root-mean-square-error (RMSE) over the bootstrap sample measures variability in conjunction with bias[42]. We performed *m* = 200 bootstrap iterations, bootstrapping at the patient level. Equations for bias and RMSE are provided in the Supplementary Material.

### 2.4 Cutoff selection

The optimum cutoff depends on the purpose for which it is being used. There is no one correct value. We choose to evaluate 4 distinct cutoffs. The first cutoff is the value optimizing Youden’s J, that is the sum of sensitivity and specificity minus one, a widely used statistical optimality criterion [43] which aims to maximise both sensitivity and specificity and treats both as equally important. The second cutoff is selected such that the percentages of observations below cutoff equals the percentages of observations with the respective outcome. We hence refer to this cutoff as the “equal proportions” cutoff. Such cutoff is particularly suited for studies aiming to estimate incidence, prevalence or risk of PD-MCI and PDD utilizing the MoCA to define a proxy outcome. While equating proportions is not a standard approach to the selection of cutoffs, it is - from a medical literature perspective - equivalent to choosing an equipercentile method for equating of scales [44]. At the equal proportions cutoff false positives equal false negatives, hence sensitivity equals positive predictive value (PPV), and specificity negative predictive value (NPV).

For screening or diagnosis of patients in clinic, the choice of cutoff carries direct implications for patients and hence the choice of a cutoff needs to take benefits and potential harms to patients into account [45]. We therefore provide a range of cutoff values alongside their respective statistical properties in the Supplementary Material. In the main manuscript we derive candidate cutoffs for screening and diagnosis following criteria from previous studies [9, 10, 15, 20] in reporting the lowest cutoff with sensitivity and NPV ≥ 80 as a potential screening cutoff, and the largest cutoff with specificity and PPV≥ 80 as a potential diagnostic cutoff. This choice is however predominantly aimed at enabling comparisons across studies. These cutoffs should not be directly incorporated into clinical practice.

## 3 Results

### 3.1 Demographics and cognitive outcomes

We analysed data from 1,094 PD and of 267 healthy control participants. In the control group the number of longitudinal observations available was on average 8.7 (SD 3.2; max 13). The number of longitudinal observations available was on average 6.9 (SD 3.3; max 13). Mean follow-up duration for the PD participants was 3.8 years (SD 3.8; max 12.1 years). Basic demographics and cognitive test scores of HC and PD participants, at baseline and longitudinally, are summarized in Supplemental Table 1.

Detailed information on the normative ranges for the neuropsychological test battery derived from the PPMI Controls data is provided in the Supplementary Material. All six tests showed an association with age unlikely due to chance (*p* < 0.001) with scores decreasing with increased age. Five out of six scores were associated with education, with those with longer education achieving higher scores on average (*p* ≤ 0.08). Only the performance in the HVLT-discrimination test did not show any indications of education dependence. The BLJ scores further differed between the odd/even versions of the test (*p* < 0.001) with higher scores achieved on the odd version.

In 881 PD patient visits (18.7%) neuropsychological testing indicated impairment in one domain, in 297 (6.3%) visits in two domains, and in 91 visits (1.9%) in all three domains tested. PD-MCI was diagnosed at at 920 (20%) patient visits, PDD at 104 (2.3%) visits (Table 1). As expected, MoCA scores are decreased and functional impairment increased with less favorable cognitive outcomes. However, while almost half of the PDD cases (47.1%) were recorded to experience moderate to severe functional impairment, only 19.8% percent of cases with impairment in all three tested domains experienced moderate to severe functional impairment (Table 1), reflecting that clinical dementia diagnoses incorporated functional information not present in the neuropsychological tests.

**Figure 1:**
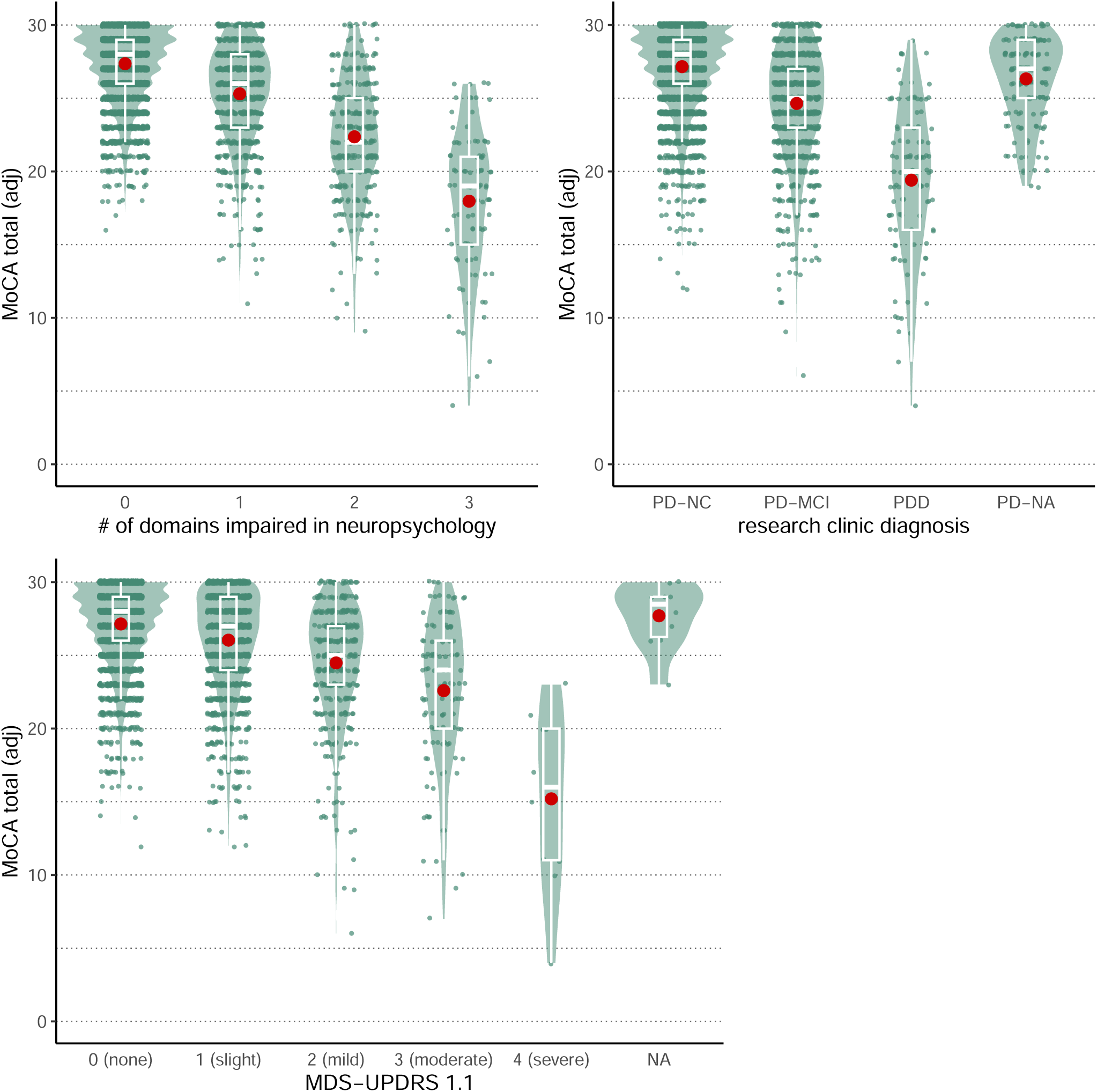
MoCA total scores (adjusted for years of education) in the PD arm of the PPMI cohort stratified by (A) number of domains impaired in neuropsychologcial testing, (B) cognitive research clinic diagnosis and (C) functional impairment as recorded in MDS-UPDRS 1 Question 1. Means, standard deviations and numbers of observations are listed in Table 1.

### 3.2 Predicting cognitive outcomes

#### 3.2.1 Neuropsychology

The education adjusted MoCA total scores discriminate patients with ≥ 1 impaired domain from those with no impairment in detailed neuropsychological testing with an AUC of 0.76, and discriminate patients with ≥ 2 impaired domains from those with one or none with an AUC of 0.86 (Table 2, Figure 2A,B). Incorporating demographics did not improve discrimination. MDS-UPDRS 1.1 or using MoCA subscores instead of the total improves discrimination slightly (Table 2, Figure 3A,B). Except for naming and abstraction all MoCA items were predictive (*p* < 0.001, Supplemental Tables 7,8). Also, all MDS-UPDRS 1.1 factor levels were predictive with *p* < 0.001 indicating that even slight functional impairment is associated with an increased probability of unfavorable performance on neuropsychological testing.

**Figure 2:**
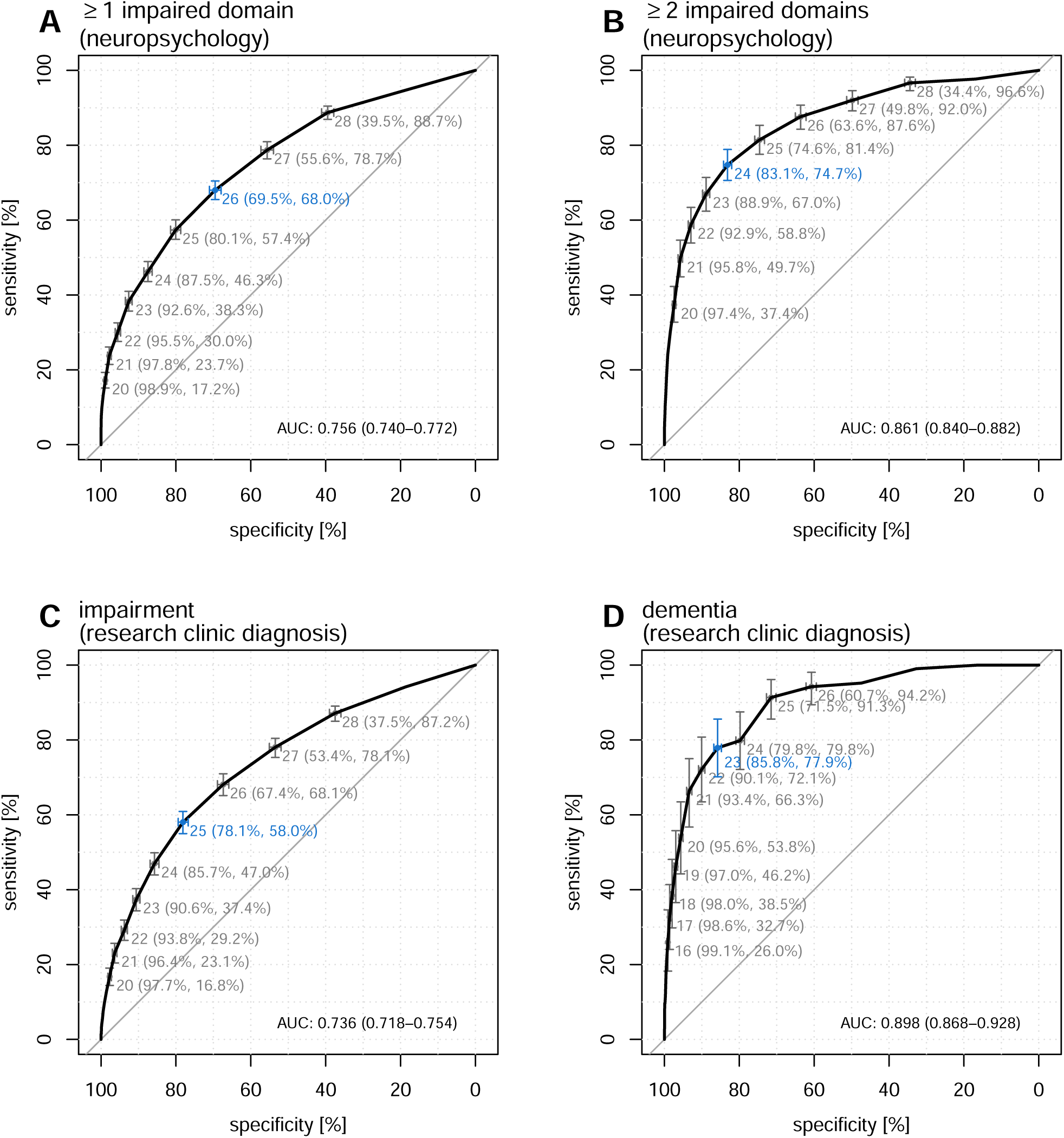
Discriminative ability of the MoCA total scores (adjusted for years of education): ROC curves and AUC’s for 4 different cognitive outcomes: (A) ≥ 1 impaired domain in neuropsychologcial testing, (B) ≥ 2 impaired domains in neuropsychologcial testing, (C) research clinic diagnosis of any impairment (PD-MCI or PDD) vs. none (PD-NC), (D) research clinic diagnosis of dementia (PDD) vs. no dementia (PD-MCI or PD-NC). The cutpoints maximizing Youden’s J = sensitivity + specificity −1 are highlighted in blue.

**Figure 3:**
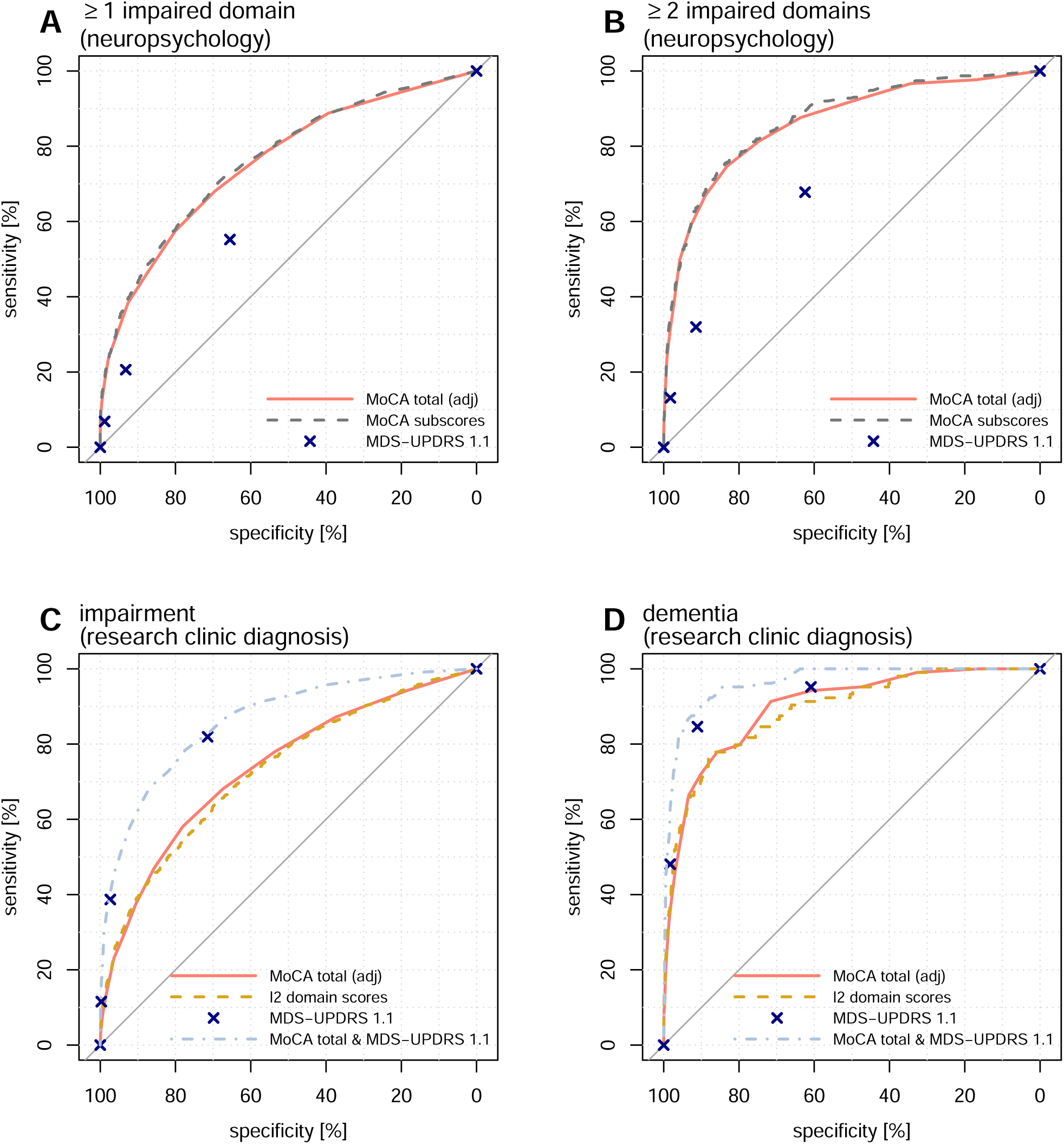
ROC curves of MoCA total scores (adjusted for years of education), MDS-UDPDRS 1 Question 1 and selected prediction models (see legends in panels) for 4 different cognitive outcomes: (A) ≥ 1 impaired domain in neuropsychologcial testing, (B) ≥ 2 impaired domains in neuropsychologcial testing, (C) research clinic diagnosis of any impairment (PD-MCI or PDD) vs. none (PD-NC), (D) research clinic diagnosis of dementia (PDD) vs. no dementia (PD-MCI or PD-NC). The corresponding AUC’s are listed in Table 2.

**Table 2:** Area under the ROC curve (AUC) of prediction models for the different cognitive outcomes utilizing different sets of covariates. Bias/Optimism correction and RMSE were estimated from *n* = 200 bootstrap iterations.

| outcome | covariates | AUC<br>(corrected) | Bias | RMSE | AUC<br>(original) |
| --- | --- | --- | --- | --- | --- |
| $\geq 1$ impaired domain<br>(neuropsychology) | MoCA total (adj) | 0.757 | -0.000 | 0.012 | 0.756 |
|  | MoCA total (raw), demographics | 0.755 | 0.004 | 0.012 | 0.759 |
|  | MoCA subscores | 0.763 | 0.001 | 0.011 | 0.764 |
|  | MoCA subscores, demographics | 0.760 | 0.005 | 0.012 | 0.765 |
|  | MDS-UPDRS 1.1 | 0.622 | -0.001 | 0.013 | 0.622 |
|  | MoCA total (adj), MDS-UPDRS 1.1 | 0.763 | 0.002 | 0.012 | 0.765 |
|  | MoCA total (raw), demogr., MDS-UPDRS 1.1 | 0.766 | 0.002 | 0.012 | 0.769 |
| $\geq 2$ impaired domains<br>(neuropsychology) | MoCA total (adj) | 0.861 | -0.000 | 0.014 | 0.861 |
|  | MoCA total (raw), demographics | 0.862 | 0.001 | 0.014 | 0.864 |
|  | MoCA subscores | 0.869 | 0.002 | 0.011 | 0.871 |
|  | MoCA subscores, demographics | 0.870 | 0.003 | 0.012 | 0.873 |
|  | MDS-UPDRS 1.1 | 0.687 | -0.002 | 0.018 | 0.685 |
|  | MoCA total (adj), MDS-UPDRS 1.1 | 0.866 | 0.001 | 0.013 | 0.867 |
|  | MoCA total (raw), demogr., MDS-UPDRS 1.1 | 0.865 | 0.004 | 0.013 | 0.869 |
| impairment<br>(research clinic diagnosis) | MoCA total (adj) | 0.738 | -0.002 | 0.014 | 0.736 |
|  | MoCA total (raw), demographics | 0.756 | 0.000 | 0.012 | 0.756 |
|  | MoCA subscores | 0.740 | 0.002 | 0.013 | 0.742 |
|  | MoCA subscores, demographics | 0.762 | 0.004 | 0.012 | 0.766 |
|  | domain scores | 0.728 | 0.001 | 0.013 | 0.729 |
|  | domain scores, demographics | 0.775 | 0.001 | 0.012 | 0.776 |
|  | MDS-UPDRS 1.1 | 0.812 | 0.000 | 0.008 | 0.812 |
|  | MoCA total (adj), MDS-UPDRS 1.1 | 0.863 | 0.000 | 0.008 | 0.863 |
|  | MoCA total (raw), demogr., MDS-UPDRS 1.1 | 0.870 | 0.001 | 0.008 | 0.871 |
| dementia<br>(research clinic diagnosis) | MoCA total (adj) | 0.897 | 0.001 | 0.015 | 0.898 |
|  | MoCA total (raw), demographics | 0.883 | 0.004 | 0.019 | 0.887 |
|  | MoCA subscores | 0.897 | 0.007 | 0.016 | 0.903 |
|  | MoCA subscores, demographics | 0.891 | 0.011 | 0.022 | 0.901 |
|  | domain scores | 0.887 | 0.002 | 0.020 | 0.889 |
|  | domain scores, demographics | 0.915 | 0.006 | 0.014 | 0.921 |
|  | MDS-UPDRS 1.1 | 0.914 | 0.003 | 0.017 | 0.917 |
|  | MoCA total (adj), MDS-UPDRS 1.1 | 0.963 | 0.001 | 0.008 | 0.964 |
|  | MoCA total (raw), demogr., MDS-UPDRS 1.1 | 0.961 | 0.004 | 0.010 | 0.964 |

Internal bootstrap validation shows no indication of overfitting for any of the models with optimism^1^ in terms of AUC estimated to be ≤ 0.002 for the majority of models and ≤ 0.005 for the most complex ones with the greatest number of covariates (Table 2). The models are overall reasonably well calibrated, but do exhibit some tendency towards too moderate estimations (calibration slopes > 1, Supplemental Tables 12, 11, Supplemental Figure 2).

Table 3 summarizes selected cutoffs for patient stratification. As no model leads to a substantial improvement over the MoCA - the maximum increase in AUC is ≤ 0.01 - we consider only cutoff on the MoCA scale (Figure 3A,B). Youden’s J is maximized at a MoCA cutpoint of ≤ 26 for stratifying ≥ 1 impaired domain; ≤ 24 for stratifying ≥ 2 impaired domains. Equal proportions cutoffs are ≤ 26 and ≤ 21, respectively, with sensitivities of 57.4% and 49.7%, meaning when equating proportions about half of cases with the respective outcome are correctly identified. Screening cutoffs are higher; diagnostic cutoffs lower.

**Table 3:**
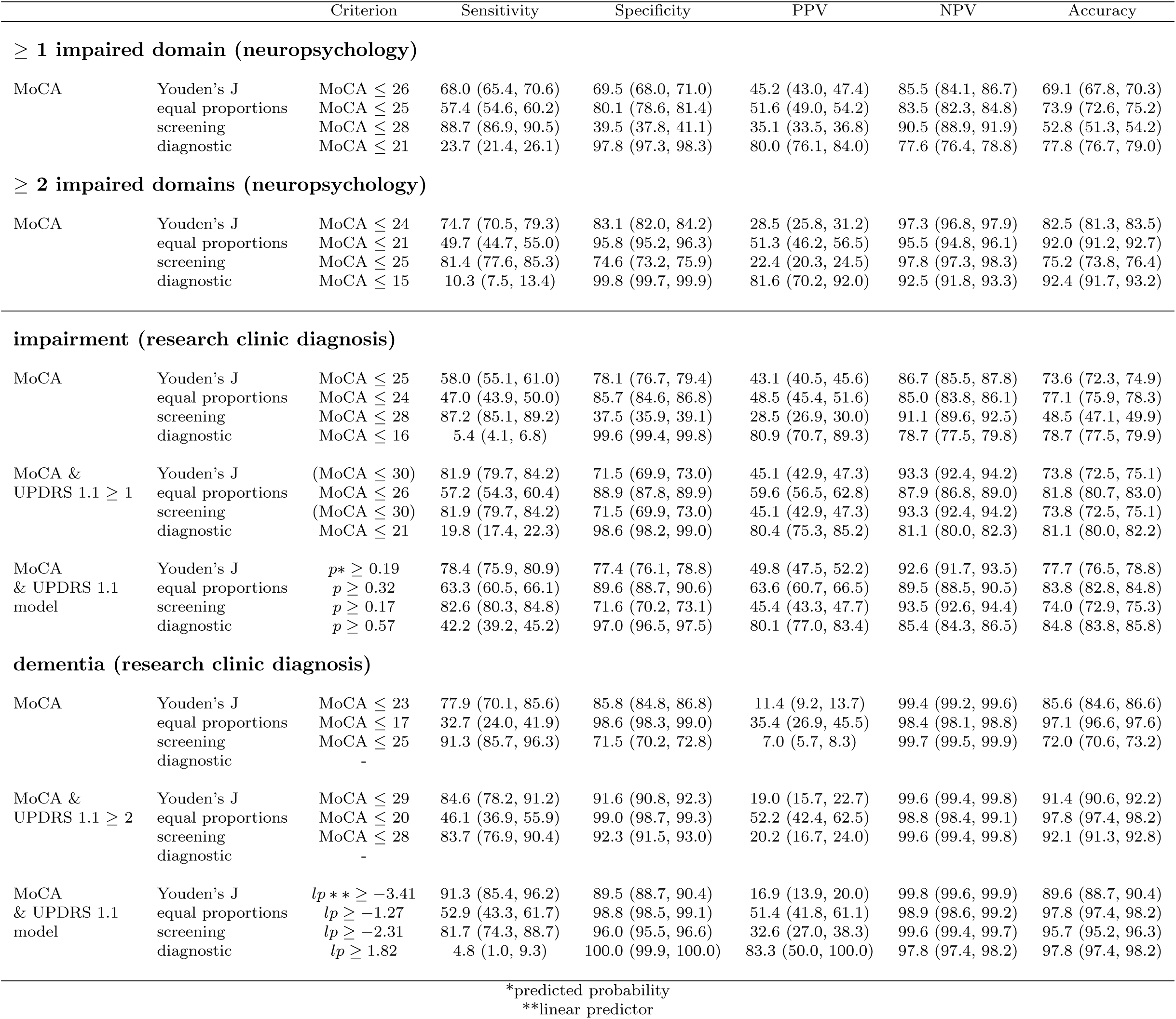
Selected cutoffs for 4 different cognitive outcomes and their respective specificity, sensitivity, positive and negative predictive values and accuracy (percentage correctly identified) with corresponding 95%-CIs.

#### 3.2.2 Research clinic diagnoses

The education adjusted MoCA total scores discriminate patients with research clinic diagnosis of any cognitive impairment (PD-MCI/PDD) from those with normal cognition with an AUC of 0.74; those with research clinic diagnosis of dementia from those with no dementia diagnosis with an AUC of 0.90 (Table 2, Figure 2C,D). Incorporating demographic covariates improves discrimination of cognitive impairment (*p* = 0.001) with age being the most relevant demographic covariate. Of two patients with the same MoCA total score, the older patient is more likely diagnosed with cognitive impairment at a research clinic visit (*p* < 0.001, Supplementary Table 9). Although this is also true for dementia diagnosis (*p* = 0.001, Supplementary Table 10), in our data set overall discrimination between dementia and no dementia is not improved by incorporating demographic covariates into the model. Also utilizing MoCA subscores instead of the totals does not improve discrimination. Interestingly, however, visuospatial, attention and orientation subscores are more strongly associated with dementia diagnosis (*p* < 0.001) than the other subscores (*p ≥* 0.15).

The greatest improvement in discrimination is achieved by adding the MDS-UPDRS 1.1 as a model covariate. This holds for both, discriminating any cognitive impairment from normal cognition (AUC=0.86) as well as discriminating dementia from no dementia (AUC=0.96). Our data even suggests, that the MDS-UPDRS 1.1 on it’s own is a superior predictor of research clinic cognitive diagnoses than the MoCA totals (Table 2, Figure 3C,D). For predicting any impairment all factor levels are relevant (*OR_M_ ≥* 5, *p* < 0.001), for predicting dementia diagnosis only MDS-UDPRS 1.1 scores of 2 or greater are predictive (*OR_M_ >* 5, *p* < 0.001) while an MDS-UPDRS 1.1 sore of 1, that is slight functional impairment, exhibited an in comparison weak association with dementia diagnosis (*OR_M_ ≈* 1.4, 0.18 *≤ p ≤* 0.39). The domain scores derived from the neuropsychological test battery were not superior compared to the MoCA in predicting research clinic diagnosis.

Internal bootstrap validation shows no indication of overfitting for any of the models with optimism in terms of AUC estimated to be ≤ 0.002 for the majority of models and ≤ 0.004 for the more complex ones. Only for the more complex models utilizing the MoCA subscores for predicting dementia diagnosis optimism is estimated to be slightly larger (0.007 to 0.011) reflecting the less powerful events per covariate ratios of these models. The models predicting any cognitive impairment show reasonable calibration. The models predicting dementia diagnosis are not reasonably calibrated (Supplemental Tables 11,12 and Figure 2). In this case model estimated probabilities should not be interpreted as such.

Youden’s J is maximized at a MoCA cutpoint of ≤ 25 for discriminating any cognitive impairment from normal cognition, and ≤ 23 for discriminating dementia from no dementia (Table 3). Equal proportions cutoffs are ≤ 24 and ≤ 17, respectively with sensitivities of 47.0% and 32.7%. Table 3 further lists the cutoffs for the best performing model which combines education adjusted MoCA totals and MDS-UPDRS 1.1. Also a simpler approach of combining a MoCA cutoff with a MDS-UPDRS 1.1 cutoff improves discrimination compared to a pure MoCA cutoff. For discriminating any cognitive impairment from none, a combination of MDS-UPDRS 1.1 ≥ 1 and MoCA ≤ 26 optimizes with respect to our equal-proportions criterion with a sensitivity of 57.2%, an increase of 10% compared to MoCA totals only. For discriminating dementia from no dementia, a combination of MDS-UPDRS 1.1 ≥ 2 and MoCA ≤ 20 optimizes with respect to the equal-proportions criterion with a sensitivity of 46.1%, an increase of about 13% compared MoCA totals only. If in favor of a slightly higher sensitivity one might alternatively choose the combination of MDS-UPDRS 1.1 ≥ 2 and MoCA ≤ 21, as the ‘true’ optimum is estimated to lie in between.

## 4 Discussion

The MoCA predicts neuropsychological testing outcomes and cognitive diagnoses with AUCs between 0.73-0.90, a range commonly regarded “acceptable” at its lower end and “excellent” at is upper end [46]. Incorporating demographic covariates or using MoCA subscores for predicting neuropsychological testing outcomes leads to a statistically significant, but small increase in AUC of ≤ 0.01 even for the most complex models. We therefore recommend applying cutoffs on original MoCA scale instead of a more complex approach. For predicting cognitive research clinic diagnosis we recommend a combination of MoCA totals and the MDS-UPDRS 1.1, which increases the AUC by *≈* 0.06 compared to MoCA totals only. In fact, the MDS-UPDRS 1.1 by itself is a better predictor of cognitive diagnosis than the MoCA. We believe that the superiority of MDS-UPDRS 1.1 functional cognitive impairment question over cognitive tests may reflect the extent of dialogue between the clinician and patient or relative/caregiver when assessing functional impairment, which may be highly salient and relevant to the clinician’s overall diagnosis. We are aware of the circularity of this part of our analysis. Clinicians making a diagnosis following the PPMI protocol are asked to evaluate functional impairment and may review cognitive test scores including the MoCA. Our findings however do suggest that the assessment of functional impairment is the dominant factor in the research clinic diagnosis and therefore, in absence of a diagnosis, a highly valuable predictor of what a diagnosis would have been. Interestingly, functional impairment was not only predictive of dementia diagnosis, but also of MCI diagnosis. The graded assessment of functional impairment in the MDS-UPDRS 1.1 allows to capture the slight to mild functional impairments that clinicians associate with an overall diagnosis of MCI.

Our finding that the domain scores derived from the neuropsychological test battery are not superior predictors of research clinic diagnosis compared to the MoCA totals may seem surprising at first. However, this is no indication that detailed testing of the in three most relevant cognitive domains in PD may be an inferior representation of a patient’s cognitive state, but rather suggests that the performance on the MoCA plays an equally great role in the research clinic diagnosis. Reviewing performance on 5 (or more) tests is more time-consuming and might require re-consideration of the respective normative ranges while the interpretation of MoCA scores is comparably straightforward.

Two further statistical remarks are to be made. The finding of better discrimination and larger AUC for the more severe cognitive outcomes is, from a statistical perspective, not surprising. Reasonable cutoffs for more severe outcomes lie in less dense regions of the statistical distribution of the MoCA totals, which reduces mis-classification risk. Secondly, we are aware that in the medical literature, adjustments of p-values for multiple comparisons are often appropriate. For our study, we deliberately chose not to do so, as the different model versions utilize common covariates and are therefore not independent.

Which cutoffs to use depends on the respective purpose. To provide suggestions for screening and diagnostic cutoffs we have followed criteria widely applied across the literature [9, 10, 16, 20]. However, we would like to emphasize that for in-clinic use with effect on patients, such as decisions on specialist referrals, counseling or treatments, situation specific decisions on suitable cutoffs need to be made taking into consideration benefits and potential harms to patients. The Supplementary Material therefore provides the discriminative properties of a wide range of cutoffs to inform individual decisions.

In pure research applications on the other hand, for instance when harmonizing data from distinct cohorts for incidence or prevalence studies, in studies identifying risk factors or developing prediction models for future cognitive decline or dementia, or for stratification of bio samples, we recommend considering the equal proportions cutoffs. It is selected such that the percentage of observations below cutoff equals the percentage of observations with the respective outcome, and holds thus an intuitive interpretation.

Given the variety of literature on PD specific MoCA cutoffs, we here only explicitly compare our findings with the two studies with the greatest sample sizes known to us. In [17] data from 106 PD patients with normal cognition, 280 with PD-MCI, and 84 with PDD was analysed and cutoffs identified by optimizing Youden’s J. For discriminating PDD from PD without dementia they found the same cutoff of ≤ 23 as the present study. The optimal cutoff to discriminate PD-MCI of ≤ 26 from [17] is one point higher than in the present study. The difference in Youden’s J between our optimal ≤ 25 cutoff and a cutoff of ≤ 26 is however small. So, all in all our findings are consistent with [17].

The study with the largest sample size we are aware of is by [20] who analyzed data from 700 PD patients with normal cognition, 706 with PD-MCI and 374 with PDD. They derive their screening and diagnostic cutoffs using the same criteria as the present study, but conclude throughout lower cutoff values - a screening cutoff of 17 for dementia for instance. The mean MoCA score in their PD-NC group was 24.5, 20.2 in the PD-MCI group and 13.4 in the PDD group, that is several points lower than in our study. Their reference, a clinical diagnosis, is based on the MDS diagnostic criteria for MCI and dementia [47, 48] including neuropsychological testing of all 5 cognitive domains evaluated using respective Italian norms. While not fully equivalent in terms of study design, the magnitude of the differences between our findings and [20] is surprising. In the original publication of the MoCA a score of ≤ 25 was described to signify mild cognitive impairment. A mean MoCA score below that threshold in a group classified as normal cognition therefore appears counterintuitive at first glance. However, [49] found a strong education dependence of the MoCA scores and in particular, a more pronounced decline with age in those who those who spent less years in education. This suggests that potentially the difference in education levels between the study population of [20] and the PPMI cohort might explain some of the differences. However, [20] reported that just like in the regression models of the present study, demographic variables, including years of education and age, were not among the key features of their decision trees.

A diagnosis made by a clinician or the results higher-order cognitive testing are solid references to validate the MoCA against, but is not a perfect representation of a patient’s cognitive state either. While diagnostic criteria are outlined in great detail in the literature [47, 48, 50], they do include passages which leave room for interpretation, such as “Impairment on neuropsychological tests may be demonstrated in several ways: performance between 1 to 2 standard deviations (SDs) below age, education, sex, and culturally appropriate norms, […]” [48]. Already a choice of a cutoff of either 1 or 1.5 SD’s below norm will lead to systematically different categorizations of patients and thus lead to differences in MoCA cutoffs optimized to resemble such categorization. Further, as we have found working with different published norms, classification depends on the choice of normative data sets for each test. We thus believe that deriving our own normative ranges for all tests utilizing data from the controls arm of the PPMI cohort and further utilizing tobit regression to handle ceiling effects of test score, where necessary, constitutes one of the strengths of our study. We evaluate all tests in a consistent manner using data from healthy control participants matched to the PD participants. The main limitation of our study and in fact any of the studies deriving MoCA cutoffs is their uncertain level of generalizability. Therefore when screening literature for guidance on suitable MoCA cutoffs either for use in clinic or for research purposes, we recommend to not only evaluate sample sizes, but also compare the population one intends to apply the cutoffs to to the population from which optimal cutoffs were derived. With the amount of literature on PD-specific MoCA cutoffs, we believe a systematic comparison and meta-analysis is warranted. Further future work should address the generalizability of MDS-UPDRS 1.1 cutoffs in comparison to the MoCA across different study populations.

We found the MDS-UPDRS 1.1 question on cognitive functional impairment - a brief patient/caregiver report - to be a better predictor of research clinic diagnoses of PD-MCI and PDD than the widely-used, clinician-applied MoCA test. This challenges the current emphasis on formal cognitive screening tools and suggests that asking patients and caregivers about real-world cognitive difficulties is a powerful, mass-available tool for cognitive screening in PD. When combined with the MoCA, this simple functional assessment achieves excellent discrimination for both any cognitive impairment (AUC 0.86) and for dementia (AUC 0.96).

## Data Availability

This analysis used data openly available from PPMI (Tier 1 data).
Data used in the preparation of this article was obtained on 2024-07-10 from the Parkinson's Progression Markers Initiative (PPMI) database (www.ppmi-info.org/access-dataspecimens/download-data), RRID:SCR_006431. For up-to-date information on the study, visit www.ppmi-info.org.

## Footnotes

1 Model optimism refers to systematic overestimation of model performance and can be induced by too small samples sizes or fitting overly complex models.

