## Supplementary Materials for "Predicting neuropsychological testing outcomes and research clinic diagnosis of MCI and dementia in Parkinson’s disease using the MoCA"

**2 Supplementary Tables & Figures** **6**

### 1 Supplementary Methods

#### 1.1 Normative ranges for HVLT, SDMT, LNS, Animals fluency and BLJ15

##### 1.1.1 Derivation using data from the healthy control participants

To maximize sample size we utilize the full longitudinal data collected from the PPMI control participants. To account for the non-independence of repeated measures collected from the same participants we utilize mixed-effect models. We regress neuropsychological test scores on age at time of assessment assessment, years of education and sex (female $\widehat{=}$ 1 ; male $\widehat{=}$ 0). The 15-item Benton Judgment of Line Orientation is additionally adjusted for form version (odd $\widehat{=}$ 1 ; even $\widehat{=}$ 0) [1].

$$y_{ij} = \underbrace{\beta_0 + \beta_{age}\ \text{age}_{ij} + \beta_{edu}\ \text{edu}_i + \beta_{female}\ \text{female}_i (+\beta_{version}\ \text{version}_i)}_{\hat{y}^{(1)}_{ij}} + \underbrace{u_{0i} + e^{(2)}_{ij}}_{e^{(1)}_{ij}} \tag{1}$$

Here $y$ represents the observed score, $i$ indexes the participant and $j$ indexes the $j$-th observation of a participant. $\hat{y}^{(1)}$ denotes the fixed effects predictions and $e^{(1)}$ the residuals of the fixed effects predictions, which are comprised of random effects $u_{0i}$ and the residuals of the fixed and random effect predictions $e^{(2)}$.

If test scores do not exhibit notable floor or ceiling effects, we fit a linear mixed-effect model with random intercept (Eq. 1). If test scores do exhibit floor or ceiling effect we fit a tobit mixed-effects model with random intercept. Details and limits set for the tobit models are provided alongside Supplementary Table 2. In the following we will omit the superscript (1) from fixed effect estimates and residuals, i.e., $\hat{y}_{ij} := \hat{y}^{(1)}_{ij}$ and $e_{ij} := e^{(1)}_{ij}$.

Applying the fitted model allows us to estimate the expected test score $\hat{y}_{ij}$ for any individual $i$ given their age, sex and years of education. Predictions are derived from fixed effects $\beta$ (see Supplementary Table 2) as

$$\hat{y}_{ij} = \beta_0 + \beta_{age}\ \text{age}_{ij} + \beta_{edu}\ \text{edu}_i + \beta_{female}\ \text{female}_i\ (+\beta_{vers}\ \text{vers}_{ij}). \tag{2}$$

The fixed effect residuals,

$$e_{ij} = y_{ij} - \hat{y}_{ij}, \tag{3}$$

represent the age, sex and education corrected test scores and indicate how much better or worse participant $i$ performs compared to what would be expected of someone at their age, sex and level of education.

To transform the fixed effect residuals $e_{ij}$ from the distinct tests onto a comparable scale, we z-scored the residuals,

$$z_{ij} = \frac{e_{ij} - \bar{e}}{\sigma_e}. \tag{4}$$

Parameters for z-scoring, i.e., mean and standard deviation, are derived from repeated sampling from the full longitudinal data set to reduce potential bias from within-participant correlation of test scores. Precisely, we repeatedly randomly draw one observation per participant, compute the standard deviation of the sample and average of the sample standard deviations and means until convergence,

$$\bar{e} = \sum_{k=1}^{m} \frac{1}{m} \bar{e}_k = \sum_{k=1}^{m} \frac{1}{m} \frac{\sum_{(i,j)\in D_k} e_{ij}}{n}, \tag{5}$$

$$\sigma_e = \sum_{k=1}^{m} \frac{1}{m} \sigma_{e_k} = \frac{1}{m} \sqrt{\frac{\sum_{(i,j)\in D_k} (e_{ij} - \bar{e}_k)^2}{n-1}}. \tag{6}$$

Here, the $2\times n$ matrix $D_k = ((1, j_1), ..., (i, j_i), ..., (n, j_n))_k$ specifies the $k$-th sample, with $(i, j_i)$ denoting the $j_i$-th observation has been drawn from participant $i$. We used $m = 1000$ samples and estimated means and standard deviations are reported in Supplementary Table 3.

The z-scored residuals of the individual tests are then summed within each cognitive domain (excluding SDMT as discussed in the main manuscript),

$$s_{exeutive} = z_{lns} + z_{animals}, \tag{7}$$

$$s_{memory} = z_{recall} + z_{discrim}, \tag{8}$$

$$s_{visuospatial} = z_{BLJ}. \tag{9}$$

Note that apart from the visuospatial domain, the z-score sums are no longer z-scored, but have a standard deviation $< 1$ due to the positive correlation of the individual test score residuals. We could repeat the z-scoring step on the domain level, but as we eventually aim to use the scores solely to categorize participants into normal cognition or impaired cognition in each domain according, we take a more direct approach and derive the 6.5%-quantiles of the domain level scores to be used as cut-offs (Supplementary Table 4). For data perfectly following a standard normal distribution the 6.5%-quantile corresponds to -1.5 standard deviations below expectation/norm. Analogous to the estimation of mean and standard deviations for z-scoring (Eqs. 5, 6), 6.5%-quantile were estimated from repeated samples of one observation per participant until convergence.

##### 1.1.2 Application to data from PD participants

Up to this point we have merely used the data from the healthy control participants to derive age, sex and education specific cut-off points to define define impairment in the three tested domains. To apply this approach to the data from the PD participants, we:

1. Apply the regression models from Eq 1 to the data collected from the PD participants ($\beta$ coefficients see Supplementary Table 2) to estimate a expected test scores given age at time of visit, sex, and years of education (and for BLJ form version utilized).

2. We then subtract the expected test score from the actual test score to derive the age, sex and education corrected test scores (Eq. 3).

3. These differences are then standardized following Eq. 4 using the test-specific means and standard deviations from Supplementary Table 3.

4. Standardized differences are then summed within each domain (Eqs. 7-9).

5. And finally we apply the cut-points/6.5%-quantiles (Supplementary Table 4) to categorize into normal cognition or impaired cognition in each domain.

#### 1.2 Prediction modeling

##### 1.2.1 Marginal effects

Due to the non-linearity of logit link function the integral over the random effects needs to be evaluated in order to generate marginal predictions [2, 3], i.e.,

$$P(Y = 1|X = x) = \int_{-\infty}^{\infty} \frac{1}{1 + \exp(-\beta_{RE,0} - \sum_{m=1}^{p} x_m \beta_{RE,m} - u)} \; f(u) \; du, \tag{10}$$

where $f(u) = \mathcal{N}(0, \sigma_u)$ denotes the density of the random effects. Simply setting $u = 0$ would lead to non-meaningful, miscalibrated predictions (Supplementary Figs. 1, 2).

However, marginal effects $\beta_{M,m}$ can be well approximated from the the respective conditional effects $\beta_{RE,m}$ as,

$$\beta_{M,m} \approx \frac{\beta_{RE,i}}{\sqrt{(c^2 \sigma_u^2 + 1)}} \quad \text{where } c = \frac{16\sqrt{3}}{15\pi}, \tag{11}$$

with $m = 0, \ldots, p$ indexing the $p$ model covariates [4, 3]. One can then proceed to estimate the probability of an outcome for a 'new' patient utilizing the marginal effects $\beta_{M,m}$, like in a standard, i.e., non-multilevel, logistic regression as

$$P(Y = 1|X = x) = \int_{-\infty}^{\infty} \frac{1}{1 + \exp(-\beta_{M,0} - \sum_{m=1}^{p} x_m \beta_{M,m})}. \tag{12}$$

Some software implementations provide the functionality of deriving meaningful marginal predictions from a multi-level logistic regression by choosing an appropriate parameter setting. In R's *lme4* package setting 're.form = NA' however disregards the random effects and hence cannot be used to obtain meaningful marginal predictions from a multi-level logistic regression (compare Supplementary Figs. 1, 2).

##### 1.2.2 Internal validation

Internal validation was carried out using non-parametric bootstrapping. The bootstrap was performed on the participant level. Using selection with replacement we randomly select participants from our original data set until we reach the same number of participants $N$ present in our original data set. We then fit our model on the bootstrap sample[1]. The model which was fitted on the bootstrap sample is then applied to the original data set. Bias (model optimism) of the AUC is estimated from the bootstrap as the mean difference in AUC between bootstrap model applied to bootstrap sample and bootstrap model applied to the original data set,

[1]Note that when re-fitting the multilevel regression, participants whose data is contained in the bootstrap sample more than once need to be assigned distinct participant id's in each draw to ensure they are treated as distinct participants and assigned distinct random effects when the model is fitted.

$$\text{AUC optimism} = \text{Bias(AUC)} = \frac{1}{m}\sum_{k=1}^{m}\Big(\text{AUC}\big(P_k(X_k)\big) - \text{AUC}\big(P_k(X)\big)\Big). \quad (13)$$

where $X$ denotes the original, full data set, $X_k$ the $k$-th bootstrap sample and $P_k$ the prediction model fitted using $k$-th bootstrap sample. The model AUC corrected for model optimism can then be derived as AUC corrected = $\text{AUC}(P(K)) - \text{Bias(AUC)}$.

The root mean square error (RMSE) of the AUC is

$$\text{RMSE(AUC)} = \sqrt{\sum_{k=1}^{m}\frac{1}{m}\Big(\text{AUC}\big(P_k(X_k)\big) - \text{AUC}\big(P_k(X)\big)\Big)^2} \quad (14)$$

and a combined measure of bias and variability[5]. We perform $m = 200$ bootstrap iterations.

##### 1.2.3 Calibration

Good model calibration is not a prerequisite for good discrimination. However, the evaluation of the calibration provides additional insights into whether the predicted probabilities can be meaningfully interpreted as such.

Firstly, we assessed calibration in a multi-level sense as the intercept and slope (fixed effects) of a multi-level logistic regression model relating linear predictor and outcome. Such calibration is, like its standard (i.e., non-multi-level) equivalent, expected to be perfect for the dataset the model was fitted to (up to numerical accuracy). Therefore assessing such calibration is only informative in the context of model validation.

Secondly, we addressed the calibration of the marginal predictions analogous to how calibration of a standard (i.e., non-multi-level) logistic regression is routinely assessed. Respective intercepts, slopes and calibration curves reveal if the predicted probabilities are meaningful as such or if a general or particular pattern of over- or underestimation is present.

Bias and RMSE of calibration slopes and intercepts were estimated using a bootstrap analogously to the internal validation of the AUC described in the section above.

### 2 Supplementary Tables & Figures

Table 1: Basic demographics and cognitive test scores in the PPMI cohort. Listed are means and standard deviations or, where applicable, percentages. Included in the table are all baseline and follow-up visits for which basic demographics (age, sex, years of education) and total scores for the six neuropsychological tests (HVLT-immediate recall, HVLT-discrimination, ANIMALS fluency, SDMT, LNS, BJL) were available/complete. The MoCA is not carried out at baseline, but at the screening visit in PPMI, hence no summary statistics are included for the MoCA at baseline and the number of longitudinal observations of the MoCA ($n$) is smaller than for the remaining variables for which we have required completeness. The two columns on the right show the relative difference between controls and PD groups, i.e., the difference in average test score between control and PD participants divided by the standard deviation of the controls group.

| | **Controls (HC)** | | **PD** | | **(HC-PD)/SD$_{HC}$** | |
|---|---|---|---|---|---|---|
| | baseline | longitudinal | baseline | longitudinal | baseline | longitudinal |
| N | 267 | 1727 | 1205 | 5863 | | |
| age | 61.69 (11.58) | 65.23 (11.60) | 62.7 (9.75) | 65.07 (9.78) | | |
| female | 37.5% | 37.8% | 38.2% | 37.9% | | |
| education years | 16.1 (3.0) | 16.1 (3.1) | 15.9 (3.5) | 15.7 (3.4) | | |
| HVLT-immediate recall | 25.77 (4.64) | 25.87 (5.40) | 24.26 (5.18) | 23.60 (5.96) | 0.33 | 0.42 |
| HVLT-discrimination | 10.27 (2.20) | 10.55 (2.18) | 9.92 (2.27) | 10.06 (2.28) | 0.16 | 0.22 |
| ANIMALS | 21.97 (5.34) | 22.21 (5.51) | 21.10 (5.59) | 20.67 (5.83) | 0.16 | 0.28 |
| SDMT | 47.11 (10.21) | 47.09 (11.37) | 41.19 (10.89) | 39.52 (12.09) | 0.58 | 0.66 |
| LNS | 10.81 (2.68) | 10.81 (2.78) | 10.36 (2.81) | 9.98 (2.99) | 0.18 | 0.30 |
| BJL | 13.11 (2.10) | 12.87 (2.22) | 12.48 (2.43) | 12.23 (2.52) | 0.30 | 0.39 |
| MoCA | - | 27.41 (2.30) | - | 26.31 (3.36) | | |
| | - | $n = 1460$ | - | $n = 4705$ | | |

Table 2: Age, education and sex-corrections of the neuropsychological scores. Listed are $\beta$ coefficients, 95%-confidence intervals and corresponding p-values. Correction is to be applied as *corrected-score* = *raw score* - *modeled score*, with *modeled score* = $\beta_0 + \beta_{age}$ age + $\beta_{edu}$ edu + $\beta_{female}$ female + ($\beta_{version}$ version), where age equals the participants age at testing, female being a binary equal to 1 if the participant is female (0 if male), edu equals the years of education of the participant and, only applicable for the BJL, version specifies the form version (odd =0, even = 1). Coefficients were estimated by fitting a mixed effect model with random intercept. Test scores with no apparent floor or ceiling effects (SDMT, LNS, BJL) were modeled with a standard linear regression. Test scores exhibiting floor or ceiling effects (HVLT, BLJ) were modeled using tobit regression. Upper and lower limits [lower, upper] set correspond to the maximum and minimum score that can be achieved in each test, that is: HVLT-discrimination [-12, 12]; HVLT-immediate recall [ , 36]; BLJ [ , 15]. Ceiling effects were negligible for the HVLT-immediate recall in that fitting a tobit model or a standard linear regression returns practically the same coefficients.

| | $\beta_0$ | $\beta_{age}$ | | $\beta_{edu}$ | | $\beta_{female}$ | |
|---|---|---|---|---|---|---|---|
| *Memory* | | | | | | | |
| HVLT-immediate recall | 30.88 (27.56, 34.20) | -0.15 (-0.18, -0.11) | $p < 0.001$ | 0.22 (0.05, 0.38) | $p = 0.01$ | 1.78 (0.76, 2.81) | $p = 0.001$ |
| HVLT-discrimination | 14.46 (12.66, 16.26) | -0.049 (-0.069, -0.029) | $p < 0.001$ | -0.004 (-0.085, 0.078) | $p = 0.93$ | 0.98 (0.46, 1.49) | $p < 0.001$ |
| *Executive function* | | | | | | | |
| SDMT | 61.00 (54.73, 67.28) | -0.37 (-0.43, -0.31) | $p < 0.001$ | 0.56 (0.24, 0.87) | $p = 0.001$ | 2.22 (0.23, 4.21) | $p = 0.029$ |
| LNS | 13.96 (12.24, 15.68) | -0.085 (-0.102, -0.069) | $p < 0.001$ | 0.14 (0.06, 0.23) | $p = 0.001$ | -0.08 (-0.62, 0.45) | $p = 0.77$ |
| Animals fluency | 25.16 (21.64, 28.68) | -0.11 (-0.15, 0.08) | $p < 0.001$ | 0.23 (0.06, 0.41) | $p = 0.008$ | 0.94 (-0.15, 2.03) | $p = 0.091$ |
| *Visuospatial function* | | | | | | | |
| BJL | 13.56 (11.79, 15.33) | -0.035 (-0.053, -0.017) | $p < 0.001$ | 0.17 (0.09, 0.26) | $p < 0.001$ | -1.45 (-1.98, -0.92) | $p < 0.001$ |
| | | $\beta_{vers}$ | | | | | |
| | | -0.51 (-0.69, -0.32) | $p < 0.001$ | | | | |

Table 3: Mean and standard deviation of the age, sex and education corrected test scores (i.e., fixed effect residuals $e$) in the healthy control group.

| test | mean $\bar{e}$ | sd $\sigma(e)$ |
|---|---|---|
| HVLT-immediate recall | -0.11 | 4.94 |
| HVLT-discrimination | -0.94 | 2.07 |
| SDMT | -0.01 | 9.04 |
| LNS | -0.03 | 2.56 |
| Animals fluency | -0.07 | 5.26 |
| BJL | -0.45 | 2.13 |

Table 4: 6.5%-quantiles of the empirical distributions of the domain sum scores used as cutoffs for impairment in the respective cognitive domain.

| | 6.5%-quantile |
|---|---|
| $s_{exeutive}$ | -1.119 |
| $s_{memory}$ | -1.351 |
| $s_{visuospatial}$ | -1.716 |

Table 5: Specificity, sensitivity, positive and negative predictive values and accuracy (percentage correctly identified) with 95%-CIs of a range of MoCA cutoff values for the different cognitive outcomes. The MoCA cutoff was applied to the MoCA total scores adjusted for years of education.

| Outcome | Cutoff | Sensitivity | Specificity | PPV | NPV | Accuracy |
|---|---|---|---|---|---|---|
| impairment | ≤ 28 | 88.7 (86.9, 90.5) | 39.5 (37.8, 41.1) | 35.1 (33.5, 36.8) | 90.5 (88.9, 91.9) | 52.8 (51.3, 54.2) |
| ≥ 1 domain | ≤ 27 | 78.7 (76.3, 81.0) | 55.6 (53.9, 57.2) | 39.6 (37.8, 41.5) | 87.6 (86.2, 88.9) | 61.9 (60.5, 63.2) |
| (neuropsychology) | ≤ 26 | 68.0 (65.4, 70.6) | 69.5 (68.0, 71.0) | 45.2 (43.0, 47.4) | 85.5 (84.1, 86.7) | 69.1 (67.8, 70.3) |
| | ≤ 25 | 57.4 (54.6, 60.2) | 80.1 (78.6, 81.4) | 51.6 (49.0, 54.2) | 83.5 (82.3, 84.8) | 73.9 (72.6, 75.2) |
| | ≤ 24 | 46.3 (43.5, 49.2) | 87.5 (86.4, 88.5) | 57.7 (54.6, 60.6) | 81.5 (80.2, 82.7) | 76.3 (75.1, 77.6) |
| | ≤ 23 | 38.3 (35.6, 41.0) | 92.6 (91.7, 93.5) | 65.7 (62.3, 69.1) | 80.2 (79.0, 81.5) | 77.9 (76.8, 79.2) |
| | ≤ 22 | 30.0 (27.5, 32.7) | 95.5 (94.8, 96.2) | 71.3 (67.5, 75.3) | 78.7 (77.5, 79.9) | 77.8 (76.7, 79.0) |
| | ≤ 21 | 23.7 (21.4, 26.1) | 97.8 (97.3, 98.3) | 80.0 (76.1, 84.0) | 77.6 (76.4, 78.8) | 77.8 (76.7, 79.0) |
| impairment | ≤ 26 | 87.6 (84.3, 90.7) | 63.6 (62.1, 65.1) | 17.8 (16.1, 19.5) | 98.3 (97.8, 98.7) | 65.6 (64.2, 67.0) |
| ≥ 2 domains | ≤ 25 | 81.4 (77.6, 85.3) | 74.6 (73.2, 75.9) | 22.4 (20.3, 24.5) | 97.8 (97.3, 98.3) | 75.2 (73.8, 76.4) |
| (neuropsychology) | ≤ 24 | 74.7 (70.5, 79.3) | 83.1 (82.0, 84.2) | 28.5 (25.8, 31.2) | 97.3 (96.8, 97.9) | 82.5 (81.3, 83.5) |
| | ≤ 23 | 67.0 (62.3, 71.9) | 88.9 (87.9, 89.8) | 35.1 (31.7, 38.6) | 96.8 (96.2, 97.3) | 87.1 (86.1, 88.0) |
| | ≤ 22 | 58.8 (53.7, 63.9) | 92.9 (92.1, 93.7) | 42.7 (38.5, 46.9) | 96.2 (95.5, 96.7) | 90.1 (89.2, 90.9) |
| | ≤ 21 | 49.7 (44.7, 55.0) | 95.8 (95.2, 96.3) | 51.3 (46.2, 56.5) | 95.5 (94.8, 96.1) | 92.0 (91.2, 92.7) |
| | ≤ 20 | 37.4 (32.5, 42.3) | 97.4 (96.9, 97.9) | 56.4 (50.0, 62.4) | 94.5 (93.9, 95.2) | 92.5 (91.7, 93.2) |
| | ≤ 19 | 30.1 (25.6, 34.8) | 98.4 (98.0, 98.8) | 63.2 (55.7, 69.9) | 94.0 (93.3, 94.7) | 92.8 (92.1, 93.5) |
| | ≤ 18 | 24.0 (19.9, 28.3) | 99.1 (98.8, 99.4) | 71.0 (62.7, 78.2) | 93.5 (92.8, 94.2) | 92.9 (92.2, 93.6) |
| | ≤ 17 | 18.3 (14.6, 22.2) | 99.4 (99.2, 99.7) | 74.0 (64.1, 82.4) | 93.1 (92.4, 93.8) | 92.7 (92.0, 93.5) |
| | ≤ 16 | 13.4 (10.1, 16.8) | 99.6 (99.4, 99.8) | 76.5 (65.7, 86.2) | 92.8 (92.0, 93.5) | 92.5 (91.8, 93.2) |
| | ≤ 15 | 10.3 (7.5, 13.4) | 99.8 (99.7, 99.9) | 81.6 (70.2, 92.0) | 92.5 (91.8, 93.3) | 92.4 (91.7, 93.2) |
| impairment | ≤ 28 | 87.2 (85.1, 89.2) | 37.5 (35.9, 39.1) | 28.5 (26.9, 30.0) | 91.1 (89.6, 92.5) | 48.5 (47.1, 49.9) |
| (clinician) | ≤ 27 | 78.1 (75.6, 80.6) | 53.4 (51.8, 55.0) | 32.4 (30.6, 34.2) | 89.5 (88.3, 90.8) | 58.9 (57.5, 60.4) |
| | ≤ 26 | 68.1 (65.4, 70.9) | 67.4 (65.8, 68.9) | 37.4 (35.2, 39.5) | 88.1 (86.9, 89.3) | 67.5 (66.2, 68.9) |
| | ≤ 25 | 58.0 (55.1, 61.0) | 78.1 (76.7, 79.4) | 43.1 (40.5, 45.6) | 86.7 (85.5, 87.8) | 73.6 (72.3, 74.9) |
| | ≤ 24 | 47.0 (43.9, 50.0) | 85.7 (84.6, 86.8) | 48.5 (45.4, 51.6) | 85.0 (83.8, 86.1) | 77.1 (75.9, 78.3) |
| | ≤ 23 | 37.4 (34.4, 40.3) | 90.6 (89.7, 91.5) | 53.2 (49.6, 56.7) | 83.5 (82.4, 84.7) | 78.8 (77.5, 79.9) |
| | ≤ 22 | 29.2 (26.5, 32.0) | 93.8 (93.0, 94.5) | 57.2 (53.1, 61.4) | 82.2 (81.1, 83.4) | 79.4 (78.4, 80.5) |
| | ≤ 21 | 23.1 (20.6, 25.7) | 96.4 (95.7, 96.9) | 64.5 (59.6, 69.4) | 81.4 (80.2, 82.6) | 80.1 (78.9, 81.2) |
| | ≤ 20 | 16.8 (14.5, 19.1) | 97.7 (97.2, 98.2) | 67.7 (61.8, 73.3) | 80.4 (79.3, 81.6) | 79.7 (78.6, 80.9) |
| | ≤ 19 | 12.9 (10.8, 15.1) | 98.5 (98.1, 98.9) | 71.6 (64.7, 77.9) | 79.8 (78.7, 81.0) | 79.5 (78.4, 80.7) |
| | ≤ 18 | 9.8 (8.0, 11.7) | 99.1 (98.8, 99.4) | 75.6 (68.0, 82.9) | 79.3 (78.2, 80.5) | 79.2 (78.1, 80.4) |
| | ≤ 17 | 7.5 (5.9, 9.1) | 99.4 (99.2, 99.7) | 79.2 (70.7, 87.1) | 79.0 (77.8, 80.2) | 79.0 (77.9, 80.2) |
| | ≤ 16 | 5.4 (4.1, 6.8) | 99.6 (99.4, 99.8) | 80.9 (70.7, 89.3) | 78.7 (77.5, 79.8) | 78.7 (77.5, 79.9) |
| dementia | ≤ 25 | 91.3 (85.7, 96.3) | 71.5 (70.2, 72.8) | 7.0 (5.7, 8.3) | 99.7 (99.5, 99.9) | 72.0 (70.6, 73.2) |
| (clinician) | ≤ 24 | 79.8 (71.7, 87.1) | 79.8 (78.7, 81.0) | 8.4 (6.8, 10.1) | 99.4 (99.2, 99.7) | 79.8 (78.7, 81.0) |
| | ≤ 23 | 77.9 (70.1, 85.6) | 85.8 (84.8, 86.8) | 11.4 (9.2, 13.7) | 99.4 (99.2, 99.6) | 85.6 (84.6, 86.6) |
| | ≤ 22 | 72.1 (63.9, 80.6) | 90.1 (89.2, 90.9) | 14.5 (11.7, 17.6) | 99.3 (99.0, 99.5) | 89.7 (88.8, 90.5) |
| | ≤ 21 | 66.3 (57.9, 75.2) | 93.4 (92.7, 94.1) | 19.0 (15.3, 23.1) | 99.2 (98.9, 99.4) | 92.8 (92.0, 93.5) |
| | ≤ 20 | 53.9 (44.3, 63.1) | 95.6 (95.0, 96.2) | 22.3 (17.5, 27.6) | 98.9 (98.5, 99.2) | 94.7 (94.0, 95.3) |
| | ≤ 19 | 46.1 (37.0, 55.6) | 97.0 (96.5, 97.5) | 26.2 (20.3, 33.0) | 98.7 (98.4, 99.0) | 95.8 (95.3, 96.4) |
| | ≤ 18 | 38.5 (28.9, 48.2) | 98.0 (97.5, 98.4) | 30.5 (23.4, 38.6) | 98.5 (98.2, 98.9) | 96.6 (96.1, 97.1) |
| | ≤ 17 | 32.7 (24.0, 41.9) | 98.6 (98.3, 99.0) | 35.4 (26.9, 45.5) | 98.4 (98.1, 98.8) | 97.1 (96.6, 97.6) |
| | ≤ 16 | 26.0 (17.8, 34.1) | 99.1 (98.8, 99.3) | 39.7 (28.8, 51.5) | 98.3 (97.9, 98.6) | 97.4 (96.9, 97.8) |
| | ≤ 15 | 21.1 (13.6, 28.8) | 99.4 (99.2, 99.6) | 44.9 (31.4, 59.2) | 98.2 (97.8, 98.6) | 97.6 (97.2, 98.0) |
| | ≤ 14 | 17.3 (10.4, 24.7) | 99.6 (99.4, 99.7) | 48.6 (32.4, 64.9) | 98.1 (97.7, 98.5) | 97.7 (97.3, 98.1) |
| | ≤ 13 | 11.5 (5.9, 17.6) | 99.7 (99.6, 99.9) | 50.0 (29.4, 69.6) | 98.0 (97.6, 98.4) | 97.7 (97.3, 98.1) |
| | ≤ 12 | 9.6 (4.2, 15.3) | 99.8 (99.7, 99.9) | 58.8 (33.3, 81.0) | 97.9 (97.5, 98.3) | 97.8 (97.4, 98.2) |
| | ≤ 11 | 9.6 (4.2, 15.3) | 99.9 (99.8, 100.0) | 71.4 (44.4, 92.9) | 97.9 (97.5, 98.3) | 97.8 (97.4, 98.2) |
| | ≤ 10 | 6.7 (2.3, 11.6) | 100.0 (99.9, 100.0) | 77.8 (44.4, 100.0) | 97.9 (97.5, 98.3) | 97.8 (97.4, 98.2) |
| | ≤ 9 | 3.9 (0.8, 8.0) | 100.0 (99.9, 100.0) | 66.7 (20.0, 100.0) | 97.8 (97.4, 98.2) | 97.8 (97.3, 98.2) |

Table 6: Specificity, sensitivity, positive and negative predictive values and accuracy (percentage correctly identified) with 95%-CIs of a range of combined MoCA and MDS-UPDRS 1.1 cutoffs for research clinic diagnosis of any impairment (vs normal cognition) and dementia (vs no dementia). The MoCA cutoff was applied to the MoCA total scores adjusted for years of education.

| Outcome | Cutoff | Sensitivity | Specificity | PPV | NPV | Accuracy |
|---|---|---|---|---|---|---|
| | MDS-UPDRS 1.1 $\geq$ 1 & | | | | | |
| impairment (clinician) | MoCA $\leq$ 30 | 81.9 (79.7, 84.2) | 71.5 (69.9, 73.0) | 45.1 (42.9, 47.3) | 93.3 (92.4, 94.2) | 73.8 (72.5, 75.1) |
| | MoCA $\leq$ 29 | 77.0 (74.4, 79.6) | 75.6 (74.2, 77.0) | 47.5 (45.2, 49.8) | 92.0 (91.1, 93.0) | 75.9 (74.7, 77.2) |
| | MoCA $\leq$ 28 | 71.8 (69.1, 74.6) | 80.4 (79.1, 81.7) | 51.1 (48.5, 53.7) | 90.9 (89.9, 91.9) | 78.5 (77.3, 79.7) |
| | MoCA $\leq$ 27 | 64.4 (61.5, 67.3) | 84.4 (83.2, 85.5) | 54.1 (51.3, 56.9) | 89.2 (88.2, 90.3) | 79.9 (78.8, 81.1) |
| | MoCA $\leq$ 26 | 57.2 (54.3, 60.4) | 88.9 (87.8, 89.9) | 59.6 (56.5, 62.8) | 87.9 (86.8, 89.0) | 81.8 (80.7, 83.0) |
| | MoCA $\leq$ 25 | 48.8 (45.8, 52.0) | 92.1 (91.2, 93.0) | 63.9 (60.5, 67.3) | 86.3 (85.2, 87.4) | 82.5 (81.4, 83.6) |
| | MoCA $\leq$ 24 | 39.6 (36.6, 42.6) | 94.7 (93.9, 95.4) | 68.2 (64.4, 71.9) | 84.6 (83.4, 85.7) | 82.5 (81.3, 83.6) |
| | MoCA $\leq$ 23 | 31.6 (28.8, 34.4) | 96.4 (95.7, 96.9) | 71.3 (67.2, 75.4) | 83.1 (82.0, 84.3) | 82.0 (80.9, 83.0) |
| | MoCA $\leq$ 22 | 25.0 (22.4, 27.7) | 97.5 (97.0, 98.0) | 74.2 (69.6, 78.8) | 82.0 (80.8, 83.1) | 81.4 (80.2, 82.5) |
| | MoCA $\leq$ 21 | 19.8 (17.4, 22.3) | 98.6 (98.2, 99.0) | 80.4 (75.3, 85.2) | 81.1 (80.0, 82.3) | 81.1 (80.0, 82.2) |
| | MDS-UPDRS 1.1 $\geq$ 1 & | | | | | |
| dementia (clinician) | MoCA $\leq$ 30 | 95.2 (90.8, 99.0) | 60.9 (59.5, 62.2) | 5.4 (4.4, 6.4) | 99.8 (99.6, 100.0) | 61.6 (60.3, 63.0) |
| | MoCA $\leq$ 29 | 95.2 (90.8, 99.0) | 65.3 (63.9, 66.7) | 6.0 (4.9, 7.2) | 99.8 (99.7, 100.0) | 66.0 (64.6, 67.3) |
| | MoCA $\leq$ 28 | 94.2 (89.7, 98.2) | 70.2 (68.9, 71.6) | 6.9 (5.7, 8.2) | 99.8 (99.6, 99.9) | 70.8 (69.5, 72.1) |
| | MoCA $\leq$ 27 | 90.4 (84.5, 95.6) | 75.0 (73.8, 76.3) | 7.8 (6.4, 9.4) | 99.7 (99.5, 99.9) | 75.4 (74.2, 76.6) |
| | MoCA $\leq$ 26 | 89.4 (83.5, 94.9) | 80.3 (79.1, 81.5) | 9.6 (7.8, 11.5) | 99.7 (99.5, 99.9) | 80.5 (79.3, 81.7) |
| | MoCA $\leq$ 25 | 86.5 (79.8, 92.9) | 84.7 (83.6, 85.7) | 11.6 (9.5, 14.1) | 99.6 (99.4, 99.8) | 84.7 (83.6, 85.7) |
| | MoCA $\leq$ 24 | 77.9 (69.7, 85.8) | 88.6 (87.6, 89.5) | 13.8 (11.1, 16.6) | 99.4 (99.2, 99.6) | 88.4 (87.4, 89.3) |
| | MoCA $\leq$ 23 | 76.0 (67.7, 84.0) | 91.7 (90.9, 92.5) | 17.6 (14.2, 21.2) | 99.4 (99.2, 99.6) | 91.3 (90.5, 92.1) |
| | MoCA $\leq$ 22 | 70.2 (62.0, 79.0) | 94.0 (93.3, 94.7) | 21.4 (17.3, 25.9) | 99.3 (99.0, 99.5) | 93.4 (92.7, 94.2) |
| | MoCA $\leq$ 21 | 64.4 (55.6, 74.0) | 95.9 (95.3, 96.5) | 26.8 (21.8, 32.6) | 99.1 (98.9, 99.4) | 95.2 (94.5, 95.8) |
| | MoCA $\leq$ 20 | 53.9 (44.9, 64.0) | 97.2 (96.7, 97.6) | 30.6 (24.4, 37.8) | 98.9 (98.6, 99.2) | 96.2 (95.6, 96.7) |
| | MoCA $\leq$ 19 | 46.1 (37.2, 56.0) | 98.0 (97.5, 98.3) | 34.5 (27.3, 42.5) | 98.7 (98.4, 99.1) | 96.8 (96.3, 97.3) |
| | MoCA $\leq$ 18 | 38.5 (30.0, 47.7) | 98.6 (98.3, 98.9) | 39.2 (30.4, 49.0) | 98.6 (98.2, 98.9) | 97.2 (96.8, 97.7) |
| | MoCA $\leq$ 17 | 32.7 (24.5, 41.9) | 99.0 (98.8, 99.3) | 44.2 (33.8, 55.2) | 98.4 (98.1, 98.8) | 97.5 (97.1, 98.0) |
| | MoCA $\leq$ 16 | 26.0 (18.3, 34.5) | 99.3 (99.0, 99.5) | 45.8 (33.9, 59.0) | 98.3 (97.9, 98.7) | 97.6 (97.2, 98.0) |
| | MoCA $\leq$ 15 | 21.1 (13.8, 29.1) | 99.5 (99.3, 99.7) | 48.9 (34.1, 63.4) | 98.2 (97.8, 98.6) | 97.7 (97.3, 98.1) |
| | MoCA $\leq$ 14 | 17.3 (10.5, 24.8) | 99.6 (99.5, 99.8) | 52.9 (35.7, 69.7) | 98.1 (97.7, 98.5) | 97.8 (97.3, 98.2) |
| | MoCA $\leq$ 13 | 11.5 (5.8, 17.9) | 99.8 (99.6, 99.9) | 52.2 (31.6, 73.1) | 98.0 (97.5, 98.4) | 97.7 (97.3, 98.2) |
| | MoCA $\leq$ 12 | 9.6 (4.3, 15.4) | 99.9 (99.7, 100.0) | 62.5 (38.5, 85.7) | 97.9 (97.5, 98.3) | 97.8 (97.4, 98.2) |
| | MoCA $\leq$ 11 | 9.6 (4.3, 15.4) | 99.9 (99.8, 100.0) | 71.4 (45.5, 93.3) | 97.9 (97.5, 98.3) | 97.8 (97.4, 98.3) |
| | MoCA $\leq$ 10 | 6.7 (2.4, 11.7) | 100.0 (99.9, 100.0) | 77.8 (50.0, 100.0) | 97.9 (97.4, 98.3) | 97.8 (97.4, 98.2) |
| | MDS-UPDRS 1.1 $\geq$ 2 & | | | | | |
| | MoCA $\leq$ 30 | 84.6 (78.2, 91.2) | 91.0 (90.2, 91.8) | 18.1 (14.9, 21.5) | 99.6 (99.4, 99.8) | 90.9 (90.1, 91.7) |
| | MoCA $\leq$ 29 | 84.6 (78.2, 91.2) | 91.6 (90.8, 92.3) | 19.0 (15.7, 22.7) | 99.6 (99.4, 99.8) | 91.4 (90.6, 92.2) |
| | MoCA $\leq$ 28 | 83.7 (76.9, 90.4) | 92.3 (91.5, 93.0) | 20.2 (16.7, 24.0) | 99.6 (99.4, 99.8) | 92.1 (91.3, 92.8) |
| | MoCA $\leq$ 27 | 79.8 (72.5, 87.3) | 93.3 (92.6, 94.0) | 21.8 (17.9, 26.1) | 99.5 (99.3, 99.7) | 93.0 (92.3, 93.7) |
| | MoCA $\leq$ 26 | 78.8 (71.4, 86.5) | 94.4 (93.8, 95.1) | 24.9 (20.4, 29.4) | 99.5 (99.3, 99.7) | 94.1 (93.4, 94.7) |
| | MoCA $\leq$ 25 | 76.9 (69.2, 84.8) | 95.5 (94.9, 96.1) | 28.5 (23.3, 33.7) | 99.4 (99.2, 99.7) | 95.1 (94.5, 95.7) |
| | MoCA $\leq$ 24 | 69.2 (60.6, 78.0) | 96.4 (95.8, 96.9) | 30.8 (24.9, 36.7) | 99.3 (99.0, 99.5) | 95.7 (95.2, 96.3) |
| | MoCA $\leq$ 23 | 67.3 (58.6, 76.2) | 97.4 (96.9, 97.8) | 37.4 (30.4, 44.2) | 99.2 (99.0, 99.5) | 96.7 (96.2, 97.2) |
| | MoCA $\leq$ 22 | 61.5 (52.5, 70.9) | 98.1 (97.7, 98.5) | 43.2 (35.4, 51.1) | 99.1 (98.8, 99.4) | 97.3 (96.8, 97.7) |
| | MoCA $\leq$ 21 | 56.7 (47.4, 66.3) | 98.6 (98.3, 99.0) | 49.2 (40.6, 58.4) | 99.0 (98.7, 99.3) | 97.7 (97.3, 98.1) |
| | MoCA $\leq$ 20 | 46.1 (36.9, 55.9) | 99.0 (98.7, 99.3) | 52.2 (42.4, 62.5) | 98.8 (98.4, 99.1) | 97.8 (97.4, 98.2) |
| | MoCA $\leq$ 19 | 39.4 (30.5, 49.1) | 99.3 (99.0, 99.5) | 56.2 (45.2, 67.7) | 98.6 (98.2, 98.9) | 97.9 (97.5, 98.3) |
| | MoCA $\leq$ 18 | 31.7 (23.2, 40.8) | 99.6 (99.4, 99.8) | 63.5 (50.9, 76.4) | 98.4 (98.1, 98.8) | 98.0 (97.6, 98.4) |
| | MoCA $\leq$ 17 | 26.9 (18.8, 35.9) | 99.7 (99.5, 99.8) | 65.1 (51.5, 78.7) | 98.3 (97.9, 98.7) | 98.0 (97.6, 98.4) |
| | MoCA $\leq$ 16 | 22.1 (14.7, 30.7) | 99.7 (99.5, 99.8) | 63.9 (48.3, 79.3) | 98.2 (97.8, 98.6) | 97.9 (97.5, 98.3) |
| | MoCA $\leq$ 15 | 19.2 (12.1, 27.3) | 99.7 (99.6, 99.9) | 62.5 (45.5, 78.6) | 98.1 (97.8, 98.5) | 97.9 (97.5, 98.3) |
| | MoCA $\leq$ 14 | 16.4 (9.6, 23.5) | 99.8 (99.6, 99.9) | 63.0 (43.5, 80.0) | 98.1 (97.7, 98.5) | 97.9 (97.5, 98.3) |
| | MoCA $\leq$ 13 | 11.5 (5.8, 18.0) | 99.8 (99.7, 99.9) | 63.2 (40.0, 83.3) | 98.0 (97.6, 98.4) | 97.8 (97.4, 98.2) |
| | MoCA $\leq$ 12 | 9.6 (4.3, 15.7) | 99.9 (99.8, 100.0) | 71.4 (44.4, 92.9) | 97.9 (97.5, 98.4) | 97.8 (97.4, 98.3) |
| | MoCA $\leq$ 11 | 9.6 (4.3, 15.7) | 99.9 (99.8, 100.0) | 71.4 (44.4, 92.9) | 97.9 (97.5, 98.4) | 97.8 (97.4, 98.3) |
| | MoCA $\leq$ 10 | 6.7 (2.4, 11.8) | 100.0 (99.9, 100.0) | 77.8 (50.0, 100.0) | 97.9 (97.4, 98.3) | 97.8 (97.4, 98.3) |

Table 7: Estimated $\beta$ coefficients and odds ratios for prediction models of impairment according to neuropsychological testing ($\geq$ 1 impaired domain). The subscript $RE$ indicates the original estimated coefficients. The subscript $M$ indicates the coefficients for the marginal predictions.

| | $\beta_{RE}$ | $OR_{RE}$ | $p$ | $\beta_M$ | $OR_M$ |
|---|---|---|---|---|---|
| **Model 1** | | | | | |
| Intercept | 8.56 (7.58, 9.53) | | <0.001 | 6.60 (5.85, 7.35) | |
| MoCA_total_adj | -0.38 (-0.41, -0.34) | 0.69 (0.66, 0.71) | <0.001 | -0.29 (-0.32, -0.26) | 0.75 (0.73, 0.77) |
| | $\sigma\_u$ =1.40 | | | | |
| **Model 2** | | | | | |
| Intercept | 7.99 (6.49, 9.49) | | <0.001 | 6.16 (5.00, 7.32) | |
| MoCA_total | -0.37 (-0.41, -0.33) | 0.69 (0.66, 0.72) | <0.001 | -0.29 (-0.32, -0.26) | 0.75 (0.73, 0.77) |
| age | 0.00 (-0.01, 0.02) | 1.00 (0.99, 1.02) | 0.61 | 0.00 (-0.01, 0.01) | 1.00 (0.99, 1.01) |
| educyrs | 0.00 (-0.04, 0.04) | 1.00 (0.97, 1.05) | 0.83 | 0.00 (-0.03, 0.03) | 1.00 (0.97, 1.03) |
| female | 0.26 (-0.01, 0.54) | 1.30 (0.99, 1.71) | 0.058 | 0.20 (-0.01, 0.41) | 1.23 (0.99, 1.51) |
| | $\sigma\_u$ =1.40 | | | | |
| **Model 3** | | | | | |
| Intercept | 9.55 (7.80, 11.31) | | <0.001 | 7.38 (6.02, 8.74) | |
| MoCA_visospatial | -0.56 (-0.66, -0.46) | 0.57 (0.52, 0.63) | <0.001 | -0.43 (-0.51, -0.35) | 0.65 (0.60, 0.70) |
| MoCA_naming | -0.05 (-0.39, 0.29) | 0.95 (0.67, 1.34) | 0.77 | -0.04 (-0.30, 0.23) | 0.96 (0.74, 1.25) |
| MoCA_attention | -0.45 (-0.57, -0.32) | 0.64 (0.56, 0.73) | <0.001 | -0.34 (-0.44, -0.25) | 0.71 (0.64, 0.78) |
| MoCA_language | -0.44 (-0.57, -0.30) | 0.65 (0.57, 0.74) | <0.001 | -0.34 (-0.44, -0.23) | 0.71 (0.64, 0.79) |
| MoCA_abstraction | -0.32 (-0.57, -0.07) | 0.73 (0.57, 0.93) | 0.012 | -0.25 (-0.44, -0.05) | 0.78 (0.65, 0.95) |
| MoCA_memory | -0.21 (-0.28, -0.15) | 0.81 (0.75, 0.86) | <0.001 | -0.17 (-0.22, -0.11) | 0.85 (0.80, 0.89) |
| MoCA_orientation | -0.62 (-0.85, -0.39) | 0.54 (0.43, 0.68) | <0.001 | -0.48 (-0.66, -0.30) | 0.62 (0.52, 0.74) |
| | $\sigma\_u$ =1.40 | | | | |
| **Model 4** | | | | | |
| Intercept | 9.35 (7.23, 11.48) | | <0.001 | 7.23 (5.58, 8.87) | |
| MoCA_visospatial | -0.56 (-0.66, -0.45) | 0.57 (0.52, 0.64) | <0.001 | -0.43 (-0.51, -0.35) | 0.65 (0.60, 0.70) |
| MoCA_naming | -0.04 (-0.38, 0.31) | 0.96 (0.68, 1.36) | 0.83 | -0.03 (-0.30, 0.24) | 0.97 (0.74, 1.27) |
| MoCA_attention | -0.44 (-0.57, -0.31) | 0.64 (0.57, 0.73) | <0.001 | -0.34 (-0.44, -0.24) | 0.71 (0.65, 0.78) |
| MoCA_language | -0.44 (-0.57, -0.30) | 0.65 (0.57, 0.74) | <0.001 | -0.34 (-0.44, -0.23) | 0.71 (0.64, 0.79) |
| MoCA_abstraction | -0.32 (-0.57, -0.06) | 0.73 (0.57, 0.94) | 0.014 | -0.24 (-0.44, -0.05) | 0.78 (0.65, 0.95) |
| MoCA_memory | -0.22 (-0.29, -0.15) | 0.80 (0.75, 0.86) | <0.001 | -0.17 (-0.22, -0.12) | 0.84 (0.80, 0.89) |
| MoCA_orientation | -0.62 (-0.85, -0.39) | 0.54 (0.43, 0.68) | <0.001 | -0.48 (-0.66, -0.30) | 0.62 (0.52, 0.74) |
| age | 0.00 (-0.01, 0.01) | 1.00 (0.99, 1.01) | 0.9 | 0.00 (-0.01, 0.01) | 1.00 (0.99, 1.01) |
| educyrs | 0.00 (-0.04, 0.04) | 1.00 (0.96, 1.04) | 0.92 | 0.00 (-0.03, 0.03) | 1.00 (0.97, 1.03) |
| female | 0.19 (-0.09, 0.46) | 1.20 (0.92, 1.58) | 0.18 | 0.14 (-0.07, 0.36) | 1.15 (0.93, 1.43) |
| | $\sigma\_u$ =1.40 | | | | |
| **Model 5** | | | | | |
| Intercept | -1.95 (-2.15, -1.74) | | <0.001 | -1.29 (-1.43, -1.16) | |
| np1cog1 | 0.82 (0.59, 1.04) | 2.26 (1.81, 2.82) | <0.001 | 0.54 (0.40, 0.69) | 1.72 (1.49, 1.99) |
| np1cog2 | 1.58 (1.24, 1.93) | 4.87 (3.44, 6.88) | <0.001 | 1.05 (0.82, 1.28) | 2.86 (2.27, 3.61) |
| np1cog3 | 2.65 (2.08, 3.23) | 14.19 (8.00, 25.17) | <0.001 | 1.76 (1.38, 2.14) | 5.83 (3.99, 8.54) |
| | $\sigma\_u$ =1.91 | | | | |
| **Model 6** | | | | | |
| Intercept | 7.51 (6.50, 8.52) | | <0.001 | 5.76 (4.99, 6.54) | |
| MoCA_total_adj | -0.35 (-0.39, -0.31) | 0.71 (0.68, 0.73) | <0.001 | -0.27 (-0.30, -0.24) | 0.76 (0.74, 0.79) |
| np1cog1 | 0.56 (0.34, 0.77) | 1.74 (1.41, 2.16) | <0.001 | 0.43 (0.26, 0.59) | 1.53 (1.30, 1.81) |
| np1cog2 | 0.99 (0.65, 1.33) | 2.69 (1.91, 3.79) | <0.001 | 0.76 (0.50, 1.02) | 2.14 (1.65, 2.78) |
| np1cog3 | 1.52 (0.94, 2.11) | 4.59 (2.56, 8.21) | <0.001 | 1.17 (0.72, 1.62) | 3.22 (2.06, 5.04) |
| | $\sigma\_u$ =1.42 | | | | |
| **Model 7** | | | | | |
| Intercept | 7.70 (6.19, 9.20) | | <0.001 | 5.96 (4.79, 7.12) | |
| MoCA_total | -0.35 (-0.39, -0.31) | 0.70 (0.68, 0.73) | <0.001 | -0.27 (-0.30, -0.24) | 0.76 (0.74, 0.79) |
| age | -0.01 (-0.02, 0.01) | 0.99 (0.98, 1.01) | 0.44 | -0.00 (-0.01, 0.01) | 1.00 (0.99, 1.01) |
| educyrs | 0.00 (-0.04, 0.04) | 1.00 (0.96, 1.04) | 0.93 | 0.00 (-0.03, 0.03) | 1.00 (0.97, 1.03) |
| female | 0.31 (0.03, 0.58) | 1.36 (1.03, 1.78) | 0.028 | 0.24 (0.03, 0.45) | 1.27 (1.03, 1.56) |
| np1cog1 | 0.57 (0.36, 0.79) | 1.77 (1.43, 2.20) | <0.001 | 0.44 (0.28, 0.61) | 1.56 (1.32, 1.84) |
| np1cog2 | 1.01 (0.67, 1.36) | 2.76 (1.95, 3.88) | <0.001 | 0.78 (0.52, 1.05) | 2.19 (1.68, 2.86) |
| np1cog3 | 1.56 (0.97, 2.14) | 4.74 (2.65, 8.49) | <0.001 | 1.20 (0.75, 1.66) | 3.33 (2.12, 5.24) |
| | $\sigma\_u$ =1.39 | | | | |

Table 8: Estimated $\beta$ coefficients and odds ratios for prediction models of impairment according to neuropsychological testing ($\geq$ 2 impaired domains). The subscript $RE$ indicates the original estimated coefficients. The subscript $M$ indicates the coefficients for the marginal predictions.

| | $\beta_{RE}$ | $OR_{RE}$ | $p$ | $\beta_M$ | $OR_M$ |
|---|---|---|---|---|---|
| **Model 1** | | | | | |
| Intercept | 9.05 (7.79, 10.31) | | <0.001 | 6.37 (5.49, 7.26) | |
| MoCA_total_adj | -0.50 (-0.55, -0.44) | 0.61 (0.58, 0.64) | <0.001 | -0.35 (-0.39, -0.31) | 0.70 (0.68, 0.73) |
| | $\sigma\_u$ =1.71 | | | | |
| **Model 2** | | | | | |
| Intercept | 8.51 (6.39, 10.64) | | <0.001 | 6.01 (4.51, 7.51) | |
| MoCA_total | -0.50 (-0.56, -0.44) | 0.61 (0.57, 0.64) | <0.001 | -0.35 (-0.39, -0.31) | 0.70 (0.67, 0.73) |
| age | -0.00 (-0.02, 0.02) | 1.00 (0.98, 1.02) | 0.76 | -0.00 (-0.02, 0.01) | 1.00 (0.98, 1.01) |
| educyrs | 0.04 (-0.01, 0.10) | 1.04 (0.99, 1.11) | 0.14 | 0.03 (-0.01, 0.07) | 1.03 (0.99, 1.07) |
| female | 0.16 (-0.27, 0.58) | 1.17 (0.77, 1.79) | 0.47 | 0.11 (-0.19, 0.41) | 1.12 (0.83, 1.51) |
| | $\sigma\_u$ =1.71 | | | | |
| **Model 3** | | | | | |
| Intercept | 10.36 (8.24, 12.47) | | <0.001 | 7.27 (5.78, 8.75) | |
| MoCA_visospatial | -0.77 (-0.91, -0.62) | 0.46 (0.40, 0.54) | <0.001 | -0.54 (-0.64, -0.44) | 0.58 (0.53, 0.65) |
| MoCA_naming | -0.54 (-0.98, -0.11) | 0.58 (0.38, 0.90) | 0.015 | -0.38 (-0.69, -0.07) | 0.68 (0.50, 0.93) |
| MoCA_attention | -0.53 (-0.70, -0.36) | 0.59 (0.50, 0.70) | <0.001 | -0.37 (-0.49, -0.25) | 0.69 (0.61, 0.78) |
| MoCA_language | -0.50 (-0.70, -0.30) | 0.61 (0.50, 0.74) | <0.001 | -0.35 (-0.49, -0.21) | 0.70 (0.61, 0.81) |
| MoCA_abstraction | -0.41 (-0.75, -0.06) | 0.67 (0.47, 0.94) | 0.02 | -0.29 (-0.53, -0.05) | 0.75 (0.59, 0.96) |
| MoCA_memory | -0.23 (-0.34, -0.13) | 0.79 (0.71, 0.88) | <0.001 | -0.16 (-0.24, -0.09) | 0.85 (0.79, 0.92) |
| MoCA_orientation | -0.68 (-0.95, -0.40) | 0.51 (0.39, 0.67) | <0.001 | -0.47 (-0.67, -0.28) | 0.62 (0.51, 0.75) |
| | $\sigma\_u$ =1.73 | | | | |
| **Model 4** | | | | | |
| Intercept | 10.37 (7.65, 13.08) | | <0.001 | 7.39 (5.45, 9.32) | |
| MoCA_visospatial | -0.78 (-0.92, -0.63) | 0.46 (0.40, 0.53) | <0.001 | -0.55 (-0.66, -0.45) | 0.58 (0.52, 0.64) |
| MoCA_naming | -0.56 (-0.99, -0.12) | 0.57 (0.37, 0.89) | 0.012 | -0.40 (-0.71, -0.09) | 0.67 (0.49, 0.92) |
| MoCA_attention | -0.54 (-0.70, -0.37) | 0.59 (0.50, 0.69) | <0.001 | -0.38 (-0.50, -0.26) | 0.68 (0.61, 0.77) |
| MoCA_language | -0.52 (-0.72, -0.32) | 0.59 (0.49, 0.73) | <0.001 | -0.37 (-0.51, -0.23) | 0.69 (0.60, 0.80) |
| MoCA_abstraction | -0.44 (-0.78, -0.10) | 0.64 (0.46, 0.91) | 0.012 | -0.31 (-0.56, -0.07) | 0.73 (0.57, 0.93) |
| MoCA_memory | -0.24 (-0.35, -0.13) | 0.79 (0.71, 0.88) | <0.001 | -0.17 (-0.25, -0.09) | 0.84 (0.78, 0.91) |
| MoCA_orientation | -0.66 (-0.94, -0.39) | 0.51 (0.39, 0.68) | <0.001 | -0.47 (-0.67, -0.28) | 0.62 (0.51, 0.76) |
| age | -0.01 (-0.03, 0.02) | 0.99 (0.97, 1.02) | 0.59 | -0.00 (-0.02, 0.01) | 1.00 (0.98, 1.01) |
| educyrs | 0.04 (-0.02, 0.10) | 1.04 (0.98, 1.10) | 0.21 | 0.03 (-0.02, 0.07) | 1.03 (0.98, 1.07) |
| female | 0.01 (-0.42, 0.44) | 1.01 (0.66, 1.55) | 0.96 | 0.01 (-0.30, 0.31) | 1.01 (0.74, 1.37) |
| | $\sigma\_u$ =1.68 | | | | |
| **Model 5** | | | | | |
| Intercept | -6.70 (-7.75, -5.64) | | <0.001 | -2.26 (-2.61, -1.90) | |
| np1cog1 | 0.89 (0.48, 1.30) | 2.43 (1.61, 3.65) | <0.001 | 0.30 (0.16, 0.44) | 1.35 (1.17, 1.55) |
| np1cog2 | 1.73 (1.17, 2.28) | 5.63 (3.23, 9.82) | <0.001 | 0.58 (0.40, 0.77) | 1.79 (1.49, 2.16) |
| np1cog3 | 3.62 (2.82, 4.42) | 37.25 (16.73, 82.92) | <0.001 | 1.22 (0.95, 1.49) | 3.39 (2.59, 4.44) |
| | $\sigma\_u$ =4.75 | | | | |
| **Model 6** | | | | | |
| Intercept | 7.85 (6.52, 9.18) | | <0.001 | 5.51 (4.58, 6.44) | |
| MoCA_total_adj | -0.46 (-0.52, -0.41) | 0.63 (0.59, 0.66) | <0.001 | -0.33 (-0.36, -0.29) | 0.72 (0.69, 0.75) |
| np1cog1 | 0.51 (0.15, 0.87) | 1.66 (1.16, 2.39) | 0.006 | 0.36 (0.10, 0.61) | 1.43 (1.11, 1.84) |
| np1cog2 | 0.84 (0.34, 1.34) | 2.32 (1.41, 3.82) | 0.001 | 0.59 (0.24, 0.94) | 1.81 (1.27, 2.56) |
| np1cog3 | 1.65 (0.95, 2.34) | 5.19 (2.58, 10.41) | <0.001 | 1.16 (0.67, 1.64) | 3.18 (1.95, 5.18) |
| | $\sigma\_u$ =1.72 | | | | |
| **Model 7** | | | | | |
| Intercept | 8.13 (6.01, 10.25) | | <0.001 | 5.79 (4.28, 7.30) | |
| MoCA_total | -0.47 (-0.53, -0.41) | 0.63 (0.59, 0.66) | <0.001 | -0.33 (-0.37, -0.29) | 0.72 (0.69, 0.75) |
| age | -0.01 (-0.03, 0.01) | 0.99 (0.97, 1.01) | 0.23 | -0.01 (-0.02, 0.01) | 0.99 (0.98, 1.01) |
| educyrs | 0.04 (-0.02, 0.09) | 1.04 (0.98, 1.10) | 0.22 | 0.03 (-0.02, 0.07) | 1.03 (0.98, 1.07) |
| female | 0.21 (-0.21, 0.64) | 1.24 (0.81, 1.90) | 0.32 | 0.15 (-0.15, 0.46) | 1.17 (0.86, 1.58) |
| np1cog1 | 0.54 (0.18, 0.90) | 1.72 (1.20, 2.47) | 0.003 | 0.39 (0.13, 0.64) | 1.47 (1.14, 1.90) |
| np1cog2 | 0.90 (0.39, 1.40) | 2.45 (1.48, 4.04) | <0.001 | 0.64 (0.28, 1.00) | 1.89 (1.32, 2.70) |
| np1cog3 | 1.70 (1.00, 2.40) | 5.47 (2.72, 11.02) | <0.001 | 1.21 (0.71, 1.71) | 3.36 (2.04, 5.53) |
| | $\sigma\_u$ =1.67 | | | | |

Table 9: Estimated $\beta$ coefficients and odds ratios for prediction models of research clinic diagnosed impairment. The subscript $RE$ indicates the original estimated coefficients. The subscript $M$ indicates the coefficients for the marginal predictions.

| | $\beta_{RE}$ | $OR_{RE}$ | $p$ | $\beta_M$ | $OR_M$ |
|---|---|---|---|---|---|
| **Model 1** | | | | | |
| Intercept | 7.55 (6.54, 8.56) | | <0.001 | 5.50 (4.76, 6.23) | |
| MoCA_total_adj | -0.36 (-0.40, -0.32) | 0.70 (0.67, 0.72) | <0.001 | -0.26 (-0.29, -0.24) | 0.77 (0.75, 0.79) |
| | $\sigma\_u$ =1.60 | | | | |
| **Model 2** | | | | | |
| Intercept | -2.84 (-4.94, -0.75) | | 0.008 | -1.83 (-3.18, -0.48) | |
| MoCA_total | -0.32 (-0.37, -0.28) | 0.72 (0.69, 0.76) | <0.001 | -0.21 (-0.24, -0.18) | 0.81 (0.79, 0.84) |
| age | 0.13 (0.11, 0.16) | 1.14 (1.11, 1.17) | <0.001 | 0.09 (0.07, 0.10) | 1.09 (1.07, 1.11) |
| educyrs | 0.02 (-0.03, 0.07) | 1.02 (0.97, 1.08) | 0.47 | 0.01 (-0.02, 0.05) | 1.01 (0.98, 1.05) |
| female | -0.34 (-0.71, 0.04) | 0.71 (0.49, 1.04) | 0.08 | -0.22 (-0.46, 0.03) | 0.81 (0.63, 1.03) |
| | $\sigma\_u$ =2.02 | | | | |
| **Model 3** | | | | | |
| Intercept | 8.80 (6.98, 10.62) | | <0.001 | 6.35 (5.04, 7.67) | |
| MoCA_visospatial | -0.63 (-0.74, -0.52) | 0.53 (0.47, 0.59) | <0.001 | -0.46 (-0.54, -0.38) | 0.63 (0.58, 0.69) |
| MoCA_naming | 0.14 (-0.23, 0.52) | 1.16 (0.80, 1.68) | 0.45 | 0.10 (-0.16, 0.37) | 1.11 (0.85, 1.45) |
| MoCA_attention | -0.40 (-0.53, -0.26) | 0.67 (0.59, 0.77) | <0.001 | -0.29 (-0.38, -0.19) | 0.75 (0.68, 0.83) |
| MoCA_language | -0.25 (-0.40, -0.11) | 0.78 (0.67, 0.90) | 0.001 | -0.18 (-0.29, -0.08) | 0.83 (0.75, 0.93) |
| MoCA_abstraction | -0.22 (-0.49, 0.05) | 0.80 (0.62, 1.05) | 0.11 | -0.16 (-0.35, 0.04) | 0.85 (0.70, 1.04) |
| MoCA_memory | -0.19 (-0.26, -0.11) | 0.83 (0.77, 0.89) | <0.001 | -0.14 (-0.19, -0.08) | 0.87 (0.83, 0.92) |
| MoCA_orientation | -0.81 (-1.05, -0.58) | 0.44 (0.35, 0.56) | <0.001 | -0.59 (-0.76, -0.42) | 0.56 (0.47, 0.66) |
| | $\sigma\_u$ =1.63 | | | | |
| **Model 4** | | | | | |
| Intercept | -0.45 (-2.99, 2.09) | | 0.73 | -0.30 (-1.98, 1.39) | |
| MoCA_visospatial | -0.54 (-0.66, -0.43) | 0.58 (0.52, 0.65) | <0.001 | -0.36 (-0.44, -0.28) | 0.70 (0.65, 0.75) |
| MoCA_naming | 0.15 (-0.24, 0.54) | 1.16 (0.79, 1.72) | 0.45 | 0.10 (-0.16, 0.36) | 1.11 (0.85, 1.43) |
| MoCA_attention | -0.42 (-0.56, -0.27) | 0.66 (0.57, 0.76) | <0.001 | -0.28 (-0.37, -0.18) | 0.76 (0.69, 0.83) |
| MoCA_language | -0.25 (-0.41, -0.09) | 0.78 (0.66, 0.91) | 0.002 | -0.17 (-0.27, -0.06) | 0.85 (0.76, 0.94) |
| MoCA_abstraction | -0.29 (-0.58, 0.00) | 0.75 (0.56, 1.00) | 0.051 | -0.19 (-0.39, 0.00) | 0.83 (0.68, 1.00) |
| MoCA_memory | -0.13 (-0.21, -0.05) | 0.88 (0.81, 0.95) | 0.002 | -0.09 (-0.14, -0.03) | 0.92 (0.87, 0.97) |
| MoCA_orientation | -0.73 (-0.98, -0.48) | 0.48 (0.38, 0.62) | <0.001 | -0.49 (-0.65, -0.32) | 0.62 (0.52, 0.73) |
| age | 0.12 (0.10, 0.15) | 1.13 (1.10, 1.16) | <0.001 | 0.08 (0.07, 0.10) | 1.08 (1.07, 1.10) |
| educyrs | 0.01 (-0.04, 0.07) | 1.01 (0.96, 1.07) | 0.6 | 0.01 (-0.03, 0.04) | 1.01 (0.97, 1.05) |
| female | -0.42 (-0.79, -0.06) | 0.66 (0.45, 0.94) | 0.024 | -0.28 (-0.52, -0.04) | 0.76 (0.59, 0.96) |
| | $\sigma\_u$ =1.92 | | | | |
| **Model 5** | | | | | |
| Intercept | -2.58 (-2.81, -2.36) | | <0.001 | -1.82 (-1.98, -1.66) | |
| z_memory | -0.47 (-0.60, -0.34) | 0.62 (0.55, 0.71) | <0.001 | -0.33 (-0.42, -0.24) | 0.72 (0.66, 0.79) |
| z_exec | -0.94 (-1.11, -0.78) | 0.39 (0.33, 0.46) | <0.001 | -0.67 (-0.78, -0.55) | 0.51 (0.46, 0.58) |
| z_visuo | -0.29 (-0.40, -0.19) | 0.75 (0.67, 0.83) | <0.001 | -0.21 (-0.28, -0.13) | 0.81 (0.75, 0.88) |
| | $\sigma\_u$ =1.71 | | | | |
| **Model 6** | | | | | |
| Intercept | -11.01 (-12.72, -9.29) | | <0.001 | -7.42 (-8.58, -6.26) | |
| z_memory | -0.38 (-0.52, -0.25) | 0.68 (0.60, 0.78) | <0.001 | -0.26 (-0.35, -0.17) | 0.77 (0.70, 0.85) |
| z_exec | -0.96 (-1.13, -0.78) | 0.38 (0.32, 0.46) | <0.001 | -0.64 (-0.76, -0.53) | 0.52 (0.47, 0.59) |
| z_visuo | -0.28 (-0.40, -0.17) | 0.75 (0.67, 0.84) | <0.001 | -0.19 (-0.27, -0.12) | 0.83 (0.76, 0.89) |
| age | 0.14 (0.11, 0.16) | 1.15 (1.12, 1.17) | <0.001 | 0.09 (0.08, 0.11) | 1.10 (1.08, 1.11) |
| educyrs | -0.04 (-0.09, 0.01) | 0.96 (0.91, 1.01) | 0.094 | -0.03 (-0.06, 0.00) | 0.97 (0.94, 1.00) |
| female | -0.44 (-0.80, -0.09) | 0.64 (0.45, 0.92) | 0.015 | -0.30 (-0.54, -0.06) | 0.74 (0.58, 0.94) |
| | $\sigma\_u$ =1.86 | | | | |

Table 9 continued

| | $\beta_{RE}$ | $OR_{RE}$ | $p$ | $\beta_M$ | $OR_M$ |
|---|---|---|---|---|---|
| **Model 7** | | | | | |
| Intercept | -3.17 (-3.40, -2.94) | | <0.001 | -2.51 (-2.69, -2.32) | |
| np1cog1 | 2.10 (1.86, 2.34) | 8.16 (6.41, 10.38) | <0.001 | 1.66 (1.47, 1.85) | 5.26 (4.35, 6.36) |
| np1cog2 | 4.34 (3.96, 4.73) | 77.07 (52.56, 112.99) | <0.001 | 3.44 (3.13, 3.74) | 31.05 (22.94, 42.02) |
| np1cog3 | 5.71 (4.94, 6.48) | 302.14 (139.61, 653.87) | <0.001 | 4.52 (3.91, 5.13) | 91.46 (49.67, 168.42) |
| | $\sigma\_u$ =1.32 | | | | |
| **Model 8** | | | | | |
| Intercept | 4.17 (3.17, 5.17) | | <0.001 | 3.43 (2.60, 4.25) | |
| MoCA_total_adj | -0.28 (-0.32, -0.24) | 0.76 (0.73, 0.79) | <0.001 | -0.23 (-0.26, -0.19) | 0.80 (0.77, 0.82) |
| np1cog1 | 1.96 (1.71, 2.20) | 7.08 (5.55, 9.02) | <0.001 | 1.61 (1.41, 1.81) | 4.99 (4.09, 6.09) |
| np1cog2 | 4.02 (3.64, 4.41) | 55.95 (38.14, 82.07) | <0.001 | 3.31 (2.99, 3.62) | 27.26 (19.90, 37.35) |
| np1cog3 | 5.06 (4.27, 5.85) | 158.20 (71.75, 348.82) | <0.001 | 4.16 (3.51, 4.81) | 64.03 (33.44, 122.58) |
| | $\sigma\_u$ =1.18 | | | | |
| **Model 9** | | | | | |
| Intercept | -2.07 (-3.76, -0.39) | | 0.016 | -1.64 (-2.97, -0.31) | |
| MoCA_total | -0.25 (-0.29, -0.21) | 0.78 (0.75, 0.81) | <0.001 | -0.20 (-0.23, -0.16) | 0.82 (0.79, 0.85) |
| age | 0.07 (0.06, 0.09) | 1.08 (1.06, 1.10) | <0.001 | 0.06 (0.04, 0.07) | 1.06 (1.05, 1.08) |
| educyrs | 0.03 (-0.01, 0.07) | 1.03 (0.99, 1.08) | 0.19 | 0.02 (-0.01, 0.06) | 1.02 (0.99, 1.06) |
| female | -0.12 (-0.43, 0.18) | 0.88 (0.65, 1.20) | 0.42 | -0.10 (-0.34, 0.14) | 0.91 (0.71, 1.15) |
| np1cog1 | 1.93 (1.67, 2.18) | 6.86 (5.32, 8.84) | <0.001 | 1.52 (1.32, 1.72) | 4.58 (3.75, 5.60) |
| np1cog2 | 3.98 (3.59, 4.38) | 53.66 (36.06, 79.85) | <0.001 | 3.15 (2.83, 3.46) | 23.31 (17.02, 31.91) |
| np1cog3 | 4.94 (4.13, 5.76) | 140.20 (62.01, 317.02) | <0.001 | 3.91 (3.26, 4.55) | 49.80 (26.13, 94.92) |
| | $\sigma\_u$ =1.32 | | | | |

Table 10: Estimated $\beta$ coefficients and odds ratios for prediction models of research clinic diagnosed dementia. The subscript $RE$ indicates the original estimated coefficients. The subscript $M$ indicates the coefficients for the marginal predictions.

| | $\beta_{RE}$ | $OR_{RE}$ | $p$ | $\beta_M$ | $OR_M$ |
|---|---|---|---|---|---|
| **Model 1** | | | | | |
| Intercept | 6.00 (3.60, 8.41) | | <0.001 | 2.11 (1.26, 2.95) | |
| MoCA_total_adj | -0.57 (-0.68, -0.45) | 0.57 (0.50, 0.64) | <0.001 | -0.20 (-0.24, -0.16) | 0.82 (0.79, 0.85) |
| | $\sigma\_u$ =4.54 | | | | |
| **Model 2** | | | | | |
| Intercept | -13.40 (-24.38, -2.43) | | 0.017 | -3.59 (-6.52, -0.65) | |
| MoCA_total | -0.57 (-0.70, -0.43) | 0.57 (0.49, 0.65) | <0.001 | -0.15 (-0.19, -0.12) | 0.86 (0.83, 0.89) |
| age | 0.23 (0.10, 0.36) | 1.26 (1.10, 1.43) | 0.001 | 0.06 (0.03, 0.10) | 1.06 (1.03, 1.10) |
| educyrs | 0.11 (-0.10, 0.31) | 1.11 (0.91, 1.36) | 0.31 | 0.03 (-0.03, 0.08) | 1.03 (0.97, 1.09) |
| female | -0.30 (-1.93, 1.33) | 0.74 (0.15, 3.77) | 0.72 | -0.08 (-0.52, 0.36) | 0.92 (0.60, 1.43) |
| | $\sigma\_u$ =6.12 | | | | |
| **Model 3** | | | | | |
| Intercept | 5.64 (1.97, 9.32) | | 0.003 | 1.92 (0.67, 3.17) | |
| MoCA_visospatial | -0.90 (-1.22, -0.58) | 0.41 (0.29, 0.56) | <0.001 | -0.31 (-0.42, -0.20) | 0.74 (0.66, 0.82) |
| MoCA_naming | 0.17 (-0.79, 1.13) | 1.19 (0.45, 3.10) | 0.73 | 0.06 (-0.27, 0.38) | 1.06 (0.76, 1.47) |
| MoCA_attention | -0.75 (-1.09, -0.41) | 0.47 (0.33, 0.66) | <0.001 | -0.26 (-0.37, -0.14) | 0.77 (0.69, 0.87) |
| MoCA_language | -0.27 (-0.72, 0.17) | 0.76 (0.49, 1.19) | 0.23 | -0.09 (-0.24, 0.06) | 0.91 (0.78, 1.06) |
| MoCA_abstraction | -0.19 (-0.92, 0.53) | 0.82 (0.40, 1.71) | 0.6 | -0.07 (-0.31, 0.18) | 0.94 (0.73, 1.20) |
| MoCA_memory | -0.22 (-0.47, 0.03) | 0.80 (0.63, 1.03) | 0.09 | -0.07 (-0.16, 0.01) | 0.93 (0.85, 1.01) |
| MoCA_orientation | -0.93 (-1.39, -0.48) | 0.39 (0.25, 0.62) | <0.001 | -0.32 (-0.47, -0.16) | 0.73 (0.62, 0.85) |
| | $\sigma\_u$ =4.70 | | | | |
| **Model 4** | | | | | |
| Intercept | -9.12 (-18.49, 0.26) | | 0.057 | -2.73 (-5.54, 0.08) | |
| MoCA_visospatial | -0.79 (-1.14, -0.44) | 0.45 (0.32, 0.64) | <0.001 | -0.24 (-0.34, -0.13) | 0.79 (0.71, 0.88) |
| MoCA_naming | 0.13 (-0.88, 1.13) | 1.13 (0.41, 3.10) | 0.81 | 0.04 (-0.26, 0.34) | 1.04 (0.77, 1.40) |
| MoCA_attention | -0.82 (-1.19, -0.44) | 0.44 (0.30, 0.64) | <0.001 | -0.24 (-0.36, -0.13) | 0.78 (0.70, 0.88) |
| MoCA_language | -0.32 (-0.81, 0.17) | 0.73 (0.45, 1.18) | 0.2 | -0.10 (-0.24, 0.05) | 0.91 (0.79, 1.05) |
| MoCA_abstraction | -0.37 (-1.18, 0.45) | 0.69 (0.31, 1.57) | 0.38 | -0.11 (-0.35, 0.13) | 0.90 (0.70, 1.14) |
| MoCA_memory | -0.20 (-0.48, 0.07) | 0.82 (0.62, 1.08) | 0.15 | -0.06 (-0.14, 0.02) | 0.94 (0.87, 1.02) |
| MoCA_orientation | -0.91 (-1.41, -0.42) | 0.40 (0.25, 0.65) | <0.001 | -0.27 (-0.42, -0.13) | 0.76 (0.66, 0.88) |
| age | 0.19 (0.08, 0.30) | 1.21 (1.08, 1.35) | 0.001 | 0.06 (0.02, 0.09) | 1.06 (1.02, 1.09) |
| educyrs | 0.08 (-0.12, 0.27) | 1.08 (0.89, 1.31) | 0.44 | 0.02 (-0.03, 0.08) | 1.02 (0.97, 1.08) |
| female | -0.41 (-1.91, 1.08) | 0.66 (0.15, 2.94) | 0.59 | -0.12 (-0.57, 0.32) | 0.88 (0.56, 1.38) |
| | $\sigma\_u$ =5.42 | | | | |
| **Model 5** | | | | | |
| Intercept | -10.19 (-11.95, -8.44) | | <0.001 | -3.31 (-3.88, -2.74) | |
| z_memory | -0.94 (-1.31, -0.56) | 0.39 (0.27, 0.57) | <0.001 | -0.30 (-0.43, -0.18) | 0.74 (0.65, 0.83) |
| z_exec | -1.55 (-2.07, -1.03) | 0.21 (0.13, 0.36) | <0.001 | -0.50 (-0.67, -0.33) | 0.60 (0.51, 0.72) |
| z_visuo | -0.29 (-0.58, 0.01) | 0.75 (0.56, 1.01) | 0.057 | -0.09 (-0.19, 0.00) | 0.91 (0.83, 1.00) |
| | $\sigma\_u$ =4.95 | | | | |
| **Model 6** | | | | | |
| Intercept | -24.14 (-32.98, -15.30) | | <0.001 | -8.41 (-11.50, -5.33) | |
| z_memory | -0.95 (-1.34, -0.55) | 0.39 (0.26, 0.58) | <0.001 | -0.33 (-0.47, -0.19) | 0.72 (0.63, 0.83) |
| z_exec | -1.50 (-2.04, -0.97) | 0.22 (0.13, 0.38) | <0.001 | -0.52 (-0.71, -0.34) | 0.59 (0.49, 0.71) |
| z_visuo | -0.30 (-0.62, 0.01) | 0.74 (0.54, 1.01) | 0.055 | -0.11 (-0.21, 0.00) | 0.90 (0.81, 1.00) |
| age | 0.21 (0.11, 0.31) | 1.24 (1.12, 1.37) | <0.001 | 0.07 (0.04, 0.11) | 1.08 (1.04, 1.12) |
| educyrs | -0.06 (-0.21, 0.10) | 0.94 (0.81, 1.11) | 0.48 | -0.02 (-0.07, 0.04) | 0.98 (0.93, 1.04) |
| female | -0.55 (-1.84, 0.75) | 0.58 (0.16, 2.11) | 0.41 | -0.19 (-0.64, 0.26) | 0.83 (0.53, 1.30) |
| | $\sigma\_u$ =4.57 | | | | |

Table 10 continued

| | $\beta_{RE}$ | $OR_{RE}$ | $p$ | $\beta_M$ | $OR_M$ |
|---|---|---|---|---|---|
| **<u>Model 7</u>** | | | | | |
| Intercept | -9.24 (-11.07, -7.40) | | <0.001 | -3.69 (-4.42, -2.96) | |
| np1cog1 | 0.88 (-0.41, 2.17) | 2.41 (0.66, 8.76) | 0.18 | 0.35 (-0.17, 0.87) | 1.42 (0.85, 2.38) |
| np1cog2 | 4.15 (2.99, 5.31) | 63.24 (19.79, 202.04) | <0.001 | 1.66 (1.19, 2.12) | 5.24 (3.29, 8.33) |
| np1cog3 | 6.41 (5.10, 7.71) | 605.56 (164.34, 2231.43) | <0.001 | 2.56 (2.04, 3.08) | 12.91 (7.67, 21.73) |
| | $\sigma\_u$ =3.90 | | | | |
| **<u>Model 8</u>** | | | | | |
| Intercept | 1.72 (-0.42, 3.86) | | 0.11 | 1.10 (-0.27, 2.47) | |
| MoCA_total_adj | -0.37 (-0.47, -0.27) | 0.69 (0.62, 0.76) | <0.001 | -0.24 (-0.30, -0.17) | 0.79 (0.74, 0.84) |
| np1cog1 | 0.64 (-0.57, 1.86) | 1.90 (0.57, 6.39) | 0.3 | 0.41 (-0.36, 1.19) | 1.51 (0.70, 3.28) |
| np1cog2 | 3.43 (2.34, 4.52) | 30.93 (10.37, 92.27) | <0.001 | 2.20 (1.50, 2.90) | 9.02 (4.48, 18.17) |
| np1cog3 | 5.10 (3.89, 6.30) | 163.57 (49.05, 545.49) | <0.001 | 3.27 (2.49, 4.04) | 26.23 (12.12, 56.75) |
| | $\sigma\_u$ =2.04 | | | | |
| **<u>Model 9</u>** | | | | | |
| Intercept | -2.93 (-7.46, 1.61) | | 0.21 | -1.73 (-4.42, 0.95) | |
| MoCA_total | -0.38 (-0.49, -0.27) | 0.69 (0.61, 0.77) | <0.001 | -0.22 (-0.29, -0.16) | 0.80 (0.75, 0.85) |
| age | 0.05 (-0.00, 0.11) | 1.05 (1.00, 1.11) | 0.067 | 0.03 (-0.00, 0.06) | 1.03 (1.00, 1.07) |
| educyrs | 0.06 (-0.07, 0.18) | 1.06 (0.94, 1.19) | 0.37 | 0.03 (-0.04, 0.11) | 1.03 (0.96, 1.11) |
| female | 0.15 (-0.79, 1.08) | 1.16 (0.46, 2.94) | 0.76 | 0.09 (-0.47, 0.64) | 1.09 (0.63, 1.89) |
| np1cog1 | 0.55 (-0.70, 1.79) | 1.73 (0.50, 5.99) | 0.39 | 0.32 (-0.41, 1.06) | 1.38 (0.66, 2.89) |
| np1cog2 | 3.31 (2.18, 4.45) | 27.47 (8.83, 85.41) | <0.001 | 1.96 (1.29, 2.64) | 7.12 (3.64, 13.96) |
| np1cog3 | 4.93 (3.67, 6.18) | 138.01 (39.34, 484.18) | <0.001 | 2.92 (2.18, 3.66) | 18.55 (8.81, 39.02) |
| | $\sigma\_u$ =2.31 | | | | |

Table 11: Multi-level model calibration. Bias/Optimism correction and RMSE were estimated from $n = 200$ bootstrap iterations.

| | | intercept $b$ | | | | slope $a$ | | | |
|---|---|---|---|---|---|---|---|---|---|
| outcome | covariates | corrected | original | Bias | RMSE | corrected | original | Bias | RMSE |
| impairment | MoCA total (adj) | 0.01 | 0 | 0 | 0.09 | 1.01 | 1 | -0.01 | 0.05 |
| $\geq$ 1 domain | MoCA total (raw) & demographics | -0.02 | 0 | 0.02 | 0.09 | 0.99 | 1 | 0.01 | 0.05 |
| (neuropsychology) | MoCA subscores | -0.01 | 0 | 0.01 | 0.09 | 0.99 | 1 | 0.01 | 0.05 |
| | MoCA subscores, demographics | -0.02 | 0 | 0.02 | 0.08 | 0.98 | 1 | 0.02 | 0.05 |
| | MDS-UPDRS 1.1 | 0.02 | 0 | -0.02 | 0.17 | 1 | 1 | 0 | 0.1 |
| | MoCA total (adj), U 1.1 | 0 | 0 | 0.01 | 0.09 | 0.99 | 1 | 0.01 | 0.05 |
| | MoCA total (raw), dem., U1.1 | -0.02 | 0 | 0.02 | 0.09 | 0.99 | 1 | 0.01 | 0.05 |
| impairment | MoCA total (adj) | 0.01 | 0 | -0.01 | 0.2 | 1 | 1 | 0 | 0.06 |
| $\geq$ 2 domains | MoCA total (raw), demographics | 0.01 | 0 | 0 | 0.2 | 1 | 1 | 0 | 0.06 |
| (neuropsychology) | MoCA subscores | -0.04 | 0 | 0.04 | 0.21 | 0.98 | 1 | 0.02 | 0.07 |
| | MoCA subscores, demographics | -0.04 | 0 | 0.04 | 0.2 | 0.98 | 1 | 0.02 | 0.06 |
| | MDS-UPDRS 1.1 | 0.01 | 0 | 0 | 1.29 | 1.01 | 1 | -0.01 | 0.14 |
| | MoCA total (adj), U1.1 | 0 | 0 | 0 | 0.21 | 0.99 | 1 | 0.01 | 0.06 |
| | MoCA total (raw), dem., U1.1 | -0.05 | 0 | 0.05 | 0.2 | 0.99 | 1 | 0.01 | 0.06 |
| impairment | MoCA total (adj) | 0.02 | 0 | -0.02 | 0.14 | 1.01 | 1 | -0.01 | 0.06 |
| (clinician) | MoCA total (raw), demographics | 0.01 | 0 | -0.01 | 0.13 | 1.01 | 1 | -0.01 | 0.06 |
| | MoCA subscores | -0.04 | 0 | 0.04 | 0.14 | 0.98 | 1 | 0.02 | 0.06 |
| | MoCA subscores, demographics | -0.04 | 0 | 0.04 | 0.14 | 0.98 | 1 | 0.02 | 0.06 |
| | domain scores | -0.01 | 0 | 0.01 | 0.13 | 1 | 1 | 0 | 0.06 |
| | domain scores, demographics | -0.01 | 0 | 0.01 | 0.12 | 1 | 1 | 0 | 0.06 |
| | MDS-UPDRS 1.1 | -0.01 | 0 | 0.01 | 0.09 | 0.99 | 1 | 0.01 | 0.04 |
| | MoCA total (adj), U1.1 | 0 | 0 | 0.01 | 0.09 | 1 | 1 | 0 | 0.04 |
| | MoCA total (raw), dem., U1.1 | -0.01 | 0 | 0.02 | 0.09 | 0.99 | 1 | 0.01 | 0.04 |
| dementia | MoCA total (adj) | 0.05 | 0 | -0.05 | 0.81 | 1 | 1 | 0 | 0.1 |
| (clinician) | MoCA total (raw), demographics | -0.14 | 0 | 0.14 | 0.86 | 1.01 | 1 | -0.01 | 0.13 |
| | MoCA subscores | -0.38 | 0 | 0.38 | 0.87 | 0.95 | 1 | 0.06 | 0.12 |
| | MoCA subscores, demographics | -0.55 | 0 | 0.55 | 0.95 | 0.93 | 1 | 0.07 | 0.14 |
| | domain scores | -0.22 | 0 | 0.22 | 0.85 | 0.98 | 1 | 0.02 | 0.1 |
| | domain scores, demographics | -0.48 | 0 | 0.48 | 0.91 | 0.97 | 1 | 0.03 | 0.13 |
| | MDS-UPDRS 1.1 | -0.38 | -0.01 | 0.37 | 1.24 | 0.97 | 1 | 0.03 | 0.12 |
| | MoCA total (adj), U1.1 | -0.05 | 0 | 0.05 | 0.43 | 0.98 | 1 | 0.02 | 0.1 |
| | MoCA total (raw), dem., U1.1 | -0.15 | 0 | 0.15 | 0.47 | 0.99 | 1 | 0.01 | 0.1 |

Table 12: Calibration of the marginal predictions. Bias/Optimism correction and RMSE were estimated from $n = 200$ bootstrap iterations.

| | | intercept $b$ | | | | slope $a$ | | | |
|---|---|---|---|---|---|---|---|---|---|
| outcome | covariates | corrected | original | Bias | RMSE | corrected | original | Bias | RMSE |
| impairment | MoCA total (adj) | -0.02 | -0.01 | 0 | 0.05 | 1.15 | 1.14 | 0 | 0.06 |
| $\geq$ 1 domain | MoCA total (raw), demographics | -0.02 | -0.01 | 0 | 0.05 | 1.11 | 1.13 | 0.02 | 0.06 |
| (neuropsychology) | MoCA subscores | -0.01 | -0.01 | 0 | 0.05 | 1.13 | 1.14 | 0.01 | 0.06 |
| | MoCA subscores, demographics | 0 | -0.01 | 0 | 0.05 | 1.12 | 1.14 | 0.02 | 0.06 |
| | MDS-UPDRS 1.1 | -0.02 | -0.02 | -0.01 | 0.06 | 1.21 | 1.2 | -0.01 | 0.11 |
| | MoCA total (adj), U1.1 | -0.02 | -0.02 | 0 | 0.05 | 1.11 | 1.12 | 0.01 | 0.06 |
| | MoCA total (raw), dem., U1.1 | -0.02 | -0.02 | 0 | 0.05 | 1.11 | 1.12 | 0.01 | 0.06 |
| impairment | MoCA total (adj) | -0.05 | -0.06 | 0 | 0.08 | 1.19 | 1.18 | 0 | 0.06 |
| $\geq$ 2 domains | MoCA total (raw), demographics | -0.04 | -0.05 | -0.01 | 0.08 | 1.17 | 1.18 | 0.01 | 0.06 |
| (neuropsychology) | MoCA subscores | -0.05 | -0.05 | 0 | 0.08 | 1.15 | 1.17 | 0.02 | 0.06 |
| | MoCA subscores, demographics | -0.03 | -0.04 | -0.01 | 0.07 | 1.15 | 1.17 | 0.02 | 0.06 |
| | MDS-UPDRS 1.1 | -0.33 | -0.34 | -0.01 | 0.08 | 2.33 | 2.31 | -0.02 | 0.21 |
| | MoCA total (adj), U1.1 | -0.06 | -0.06 | 0 | 0.08 | 1.18 | 1.19 | 0.01 | 0.06 |
| | MoCA total (raw), dem,, U1.1 | -0.05 | -0.05 | 0 | 0.08 | 1.16 | 1.18 | 0.02 | 0.06 |
| impairment | MoCA total (adj) | 0.1 | 0.1 | 0 | 0.06 | 1.04 | 1.03 | -0.01 | 0.07 |
| (clinician) | MoCA total (raw), demographics | 0.01 | 0 | -0.01 | 0.07 | 0.86 | 0.86 | 0 | 0.05 |
| | MoCA subscores | 0.09 | 0.1 | 0.01 | 0.06 | 1.02 | 1.04 | 0.01 | 0.07 |
| | MoCA subscores, demographics | 0.02 | 0.02 | 0 | 0.07 | 0.89 | 0.91 | 0.02 | 0.06 |
| | domainscores | 0.1 | 0.11 | 0 | 0.06 | 1.03 | 1.04 | 0 | 0.07 |
| | domainscores, demographics | 0.02 | 0.03 | 0 | 0.06 | 0.92 | 0.93 | 0.01 | 0.05 |
| | MDS-UPDRS 1.1 | 0.06 | 0.06 | 0 | 0.06 | 1.11 | 1.12 | 0.01 | 0.04 |
| | MoCA total (adj), U1.1 | 0.05 | 0.04 | 0 | 0.06 | 1.05 | 1.06 | 0.01 | 0.04 |
| | MoCA total (raw), dem., U1.1 | 0.03 | 0.02 | 0 | 0.06 | 1.02 | 1.03 | 0.01 | 0.04 |
| dementia | MoCA total (adj) | -0.84 | -0.86 | -0.02 | 0.14 | 1.82 | 1.82 | 0 | 0.11 |
| (clinician) | MoCA total (raw), demographics | -0.98 | -1.01 | -0.03 | 0.13 | 1.8 | 1.84 | 0.04 | 0.14 |
| | MoCA subscores | -0.9 | -0.92 | -0.01 | 0.14 | 1.81 | 1.87 | 0.05 | 0.13 |
| | MoCA subscores, demographics | -0.92 | -0.93 | -0.01 | 0.12 | 1.75 | 1.84 | 0.08 | 0.17 |
| | domainscores | -0.9 | -0.91 | -0.01 | 0.14 | 2.14 | 2.15 | 0.02 | 0.16 |
| | domainscores, demographics | -0.67 | -0.67 | 0 | 0.14 | 1.78 | 1.85 | 0.07 | 0.14 |
| | MDS-UPDRS 1.1 | -0.60 | -0.62 | -0.02 | 0.14 | 2.13 | 2.13 | 0. | 0.14 |
| | MoCA total (adj), U1.1 | -0.12 | -0.13 | -0.01 | 0.14 | 1.26 | 1.29 | 0.02 | 0.09 |
| | MoCA total (raw), dem., U1.1 | -0.16 | -0.19 | -0.03 | 0.15 | 1.29 | 1.32 | 0.04 | 0.09 |

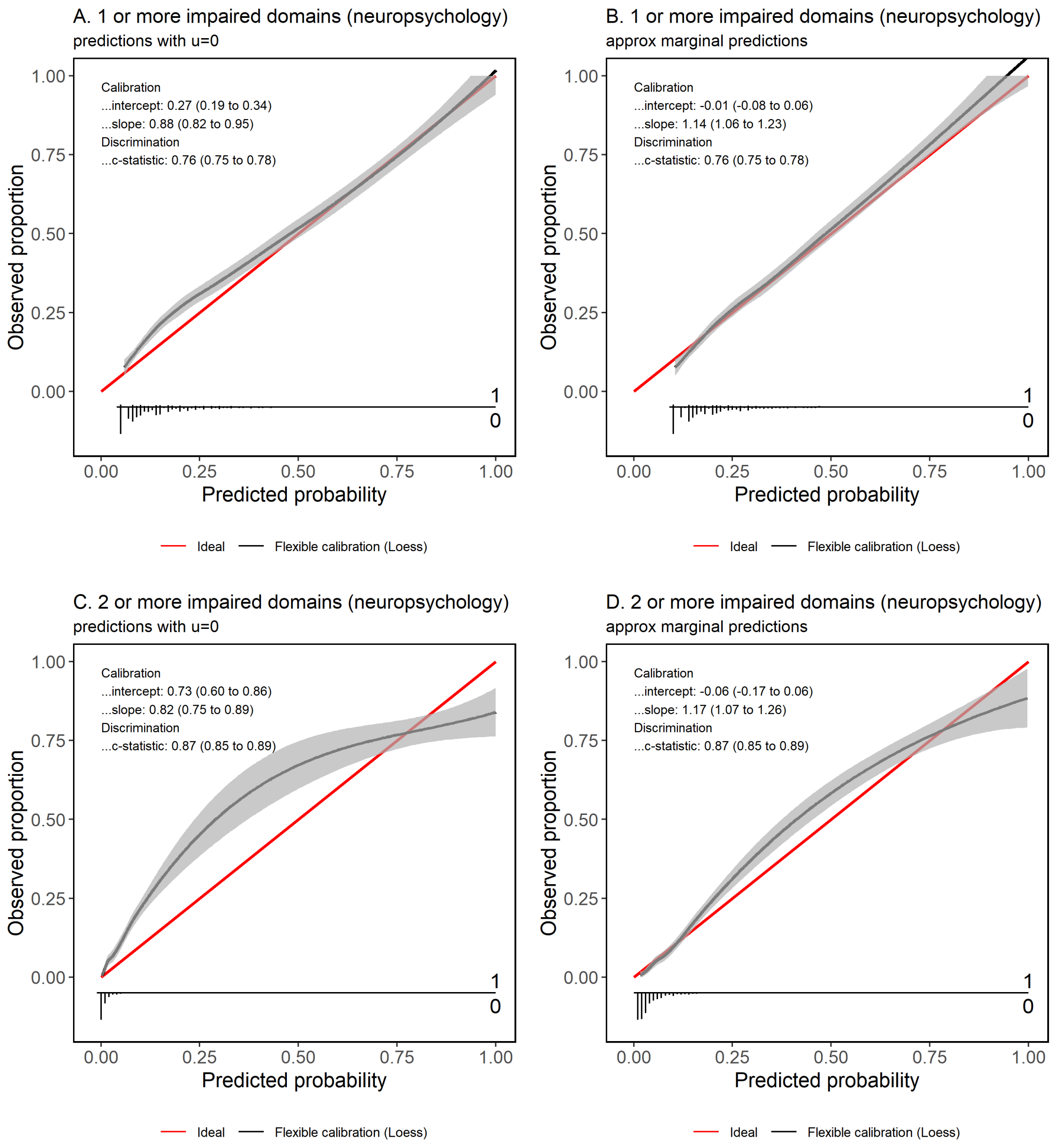

Figure 1: Calibration curves for the models predicting impairment according to neuropsychological testing from MoCA subscores. Left: Predictions using $\beta_{RE}$ and setting random effect $u = 0$. Right: Approximated marginal predictions.

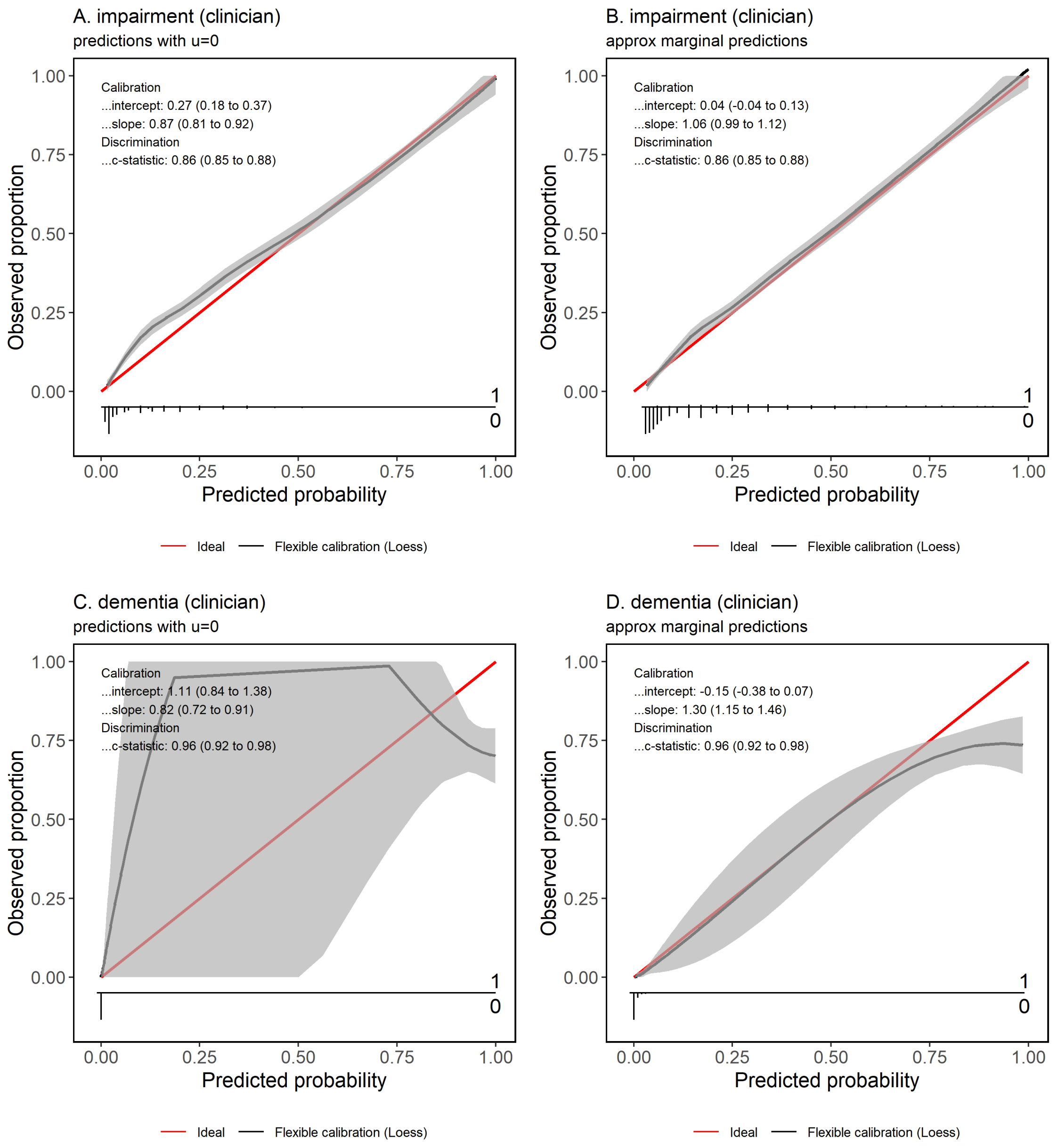

Figure 2: Calibration curves for the models predicting research clinic diagnoses from MoCA totals (adj) and MDS-UPDRS 1.1. Left: Predictions using $\beta_{RE}$ and setting random effect $u = 0$. Right: Approximated marginal predictions.
